# Viral respiratory epidemic modelling of societal segregation based on vaccination status

**DOI:** 10.1101/2022.08.21.22279035

**Authors:** Joseph Hickey, Denis G. Rancourt

## Abstract

**Background:** Societal segregation of unvaccinated people from public spaces has been a novel and controversial COVID-era public health practice in many countries. Models exploring potential consequences of vaccination-status-based segregation have not considered how segregation influences the contact frequencies in the segregated groups. We systematically investigate implementing effects of segregation on population-specific contact frequencies and show this critically determines the predicted epidemiological outcomes, focusing on the attack rates in the vaccinated and unvaccinated populations and the share of infections among vaccinated people that were due to contacts with infectious unvaccinated people.

**Methods:** We describe a susceptible-infectious-recovered (SIR) two-population model for vaccinated and unvaccinated groups of individuals that transmit an infectious disease by person-to-person contact. The degree of segregation of the two groups, ranging from zero to complete segregation, is implemented using the like-to-like mixing approach developed for sexually-transmitted diseases, adapted for presumed SARS-CoV-2 transmission. We allow the contact frequencies for individuals in the two groups to be different and depend, with variable strength, on the degree of segregation.

**Results:** Segregation can either increase or decrease the attack rate among the vaccinated, depending on the type of segregation (isolating or compounding), and the contagiousness of the disease. For diseases with low contagiousness, segregation can cause an attack rate in the vaccinated, which does not occur without segregation.

**Interpretation:** There is no predicted blanket epidemiological advantage to segregation, either for the vaccinated or the unvaccinated. Negative epidemiological consequences can occur for both groups.

## Introduction

Models can be used to investigate infectious disease dynamics under different hypotheses about the characteristics of a disease and the effects of health policy. In this endeavour, there are advantages to working with simplest-possible but sufficiently realistic models [1,2], where one should exclude simple models that are not sufficiently realistic for the intended application, either because of their structure or because of incorrect assumptions about the underlying mechanisms. Following this approach, researchers have extended the foundational simple susceptible-infectious-recovered (SIR)-type model to explore diseases with birth and death dynamics, maternal- or vaccine-derived immunity, latency of infection, patterns of contact mixing between different societal groups, and so on [3-7], and to study the effect of isolating vulnerable individuals from the general population during a pandemic, in the absence of vaccination [8].

Recently, SIR models of epidemic dynamics have been implemented with two interacting societal groups (vaccinated and unvaccinated) to examine epidemic outcomes for variable degrees of interaction between the two groups, including whether the unvaccinated put the vaccinated unduly or disproportionately at risk, using epidemiological parameters intended to be representative of SARS-CoV-2 [9-12]. These prior implementations regarding groups differentiated by vaccination status take the contact frequencies of the majority and socially-excluded groups to be equal and held constant, irrespective of the degree of segregation (or exclusion or “like-to-like mixing”), which is not realistic.

Here, we implement population-specific contact frequencies that can be different for the two groups and that can either increase or decrease with increasing segregation. This is necessary because, for example, in many actual regulatory policies the excluded unvaccinated group is barred from public venues or services where people gather and from public transport where people are in close proximity for various durations. In general, the contact frequency of the excluded group decreases with increasing segregation if isolation is in effect, and increases with increasing segregation if the excluded individuals are crowded together. Implementing this essential model feature gives rise to more complex behaviour of the attack rates in the vaccinated and unvaccinated populations (*A*_*v*_ and *A*_*u*_, respectively), which can increase or decrease, or rise to a maximum before decreasing, as the two groups are increasingly segregated. This is also true for the share (*B*_*v*_) of infections among vaccinated people that are due to contacts with infectious unvaccinated people.

This article was previously posted to the medRxiv preprint server on August 23, 2022, and revised versions were posted to medRxiv on November 28, 2022; July 6, 2023; and July 19, 2023.

## Methods

### Model Design

We adopt the standard SIR framework in a structure with two sub-populations. If a susceptible person (S) comes into contact with an infectious person (I), the susceptible person can become infectious, and infectious people eventually recover (R) and become permanently immune.

We divide the population into two groups: vaccinated and unvaccinated. Vaccination is “all or nothing”, such that a proportion VE of the vaccinated population is immune (is in the R state from the outset of the simulation), where the parameter VE represents vaccine efficacy. The model also includes a natural immunity parameter, NI, equal to the proportion of unvaccinated that are immune from the outset due to previous infection [9].

The model parameter η controls the degree of segregation of vaccinated and unvaccinated people. When η = 0, there is no segregation, and the two groups mix randomly. When η = 1, there is complete segregation, such that vaccinated only come into contact with other vaccinated, and unvaccinated only come into contact with other unvaccinated.

The parameter η follows from Garnett and Anderson [1], who modeled sexually-transmitted disease spread in a population divided into groups with different frequencies of sexual contacts. They take the contact frequency to be a constant characteristic of the individuals within a group. However, contact frequency is not generally and solely an intrinsic individual characteristic [13].

In our model, the population-specific contact frequencies of the vaccinated and unvaccinated individuals (*c*_*v*_ and *c*_*u*_, respectively) can increase, decrease, or remain constant as the two groups are segregated. We implement a new approach to achieve this: we keep the first two terms in Taylor expansions of *c*_*v*_ and *c*_*u*_ versus η (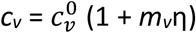 and 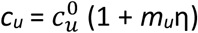; see Eqs. A3 in Appendix A1). Thus, *m*_*v*_ and *m*_*u*_ determine the degree of increase or decrease of the contact frequency in either group, as η is increased.

For example, when *m*_*u*_ < 0, as segregation is increased, the contact frequency of unvaccinated people decreases. This corresponds to segregation policy that excludes unvaccinated people from public spaces such as restaurants, cinemas, workplaces, airplanes, trains, etc. [14-17]. Conversely, *m*_*u*_ > 0 corresponds to segregation policy that increases contacts between unvaccinated people; for example, by requiring returning unvaccinated travelers to stay in designated facilities [18-20].

In principle, the vaccinated and unvaccinated contact frequencies may be different even when the two groups are completely unsegregated. The unsegregated (η = 0) contact frequencies are set by the parameters 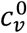 and 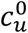. Similarly, the probability that contact between a susceptible and infectious person results in transmission is β_*v*_ (β_*u*_) for a susceptible vaccinated (unvaccinated) person and the rates of recovery from infection for the vaccinated and unvaccinated individuals are γ_*v*_ and γ_*u*_, respectively.

There are thus two “β parameters”, two “*c* parameters” and two “γ parameters” in our model. Since each β parameter always occurs as part of a product with its respective *c* parameter, the β parameters can freely be set equal to 1. We set β_*v*_ = β_*u*_ = 1 in this paper, without any loss of generality. This implies that, by definition (since β = 1), the contact frequencies “c” in our model are conceptually for contacts that are of sufficiently close proximity and long duration that an infection is guaranteed to occur when a susceptible and an infectious person meet. For a more contagious virus, more of an individual’s contacts are long and close enough that transmission would be guaranteed, corresponding to higher 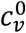 and 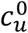.

The model of Fisman et al. [9] is the special case of our model with 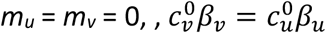 and *γ*_*v*_ = *γ*_*u*_, in which case the equal contact frequencies of both vaccinated and unvaccinated remain constant regardless of the level of segregation. Such an implementation does not represent how segregation has been applied during the COVID era in Canada and many countries [14-17,21], since unvaccinated people were excluded from public spaces while vaccinated people were allowed access, thus changing venues and opportunities for contact as segregation is imposed.

Throughout this paper, “contact frequency” refers to frequency of infectious contacts, since the probability of infection per infectious-susceptible contact is set equal to 1 without loss of generality (see Appendix 1).

### Model Parameterization

The parameters of our model are listed in Table 1; calculated quantities in Table 2. Technical details of the model are in Appendix 1.

**Table 1:** Model parameters.

| Parameter description | Symbol | Typical value | Bound |
| --- | --- | --- | --- |
| Degree of segregation of vaccinated and unvaccinated groups | $\eta$ | (varied) | 0 to 1 |
| Contact frequency of vaccinated people when $\eta=0$ | $c_v^0$ | 300 contacts/yr | $\geq 0$ |
| Contact frequency of unvaccinated people when $\eta=0$ | $c_u^0$ | 300 contacts/yr | $\geq 0$ |
| Probability of transmission per contact between a susceptible vaccinated person and an infectious person | $\beta_v$ | 1 | 0 to 1 |
| Probability of transmission per contact between a susceptible unvaccinated person and an infectious person | $\beta_u$ | 1 | 0 to 1 |
| Degree of increase ($m_v > 0$) or decrease ($m_v < 0$) of vaccinated contact frequency as a function of $\eta$ | $m_v$ | 0 | $\geq -1$ |
| Degree of increase ($m_u > 0$) or decrease ($m_u < 0$) of unvaccinated contact frequency as a function of $\eta$ | $m_u$ | (varied) | $\geq -1$ |
| Rate of recovery from infection of a vaccinated person (per year) | $\gamma_v$ | $73 \text{ yr}^{-1}$ | $\geq 0$ |
| Rate of recovery from infection of an unvaccinated person (per year) | $\gamma_u$ | $73 \text{ yr}^{-1}$ | $\geq 0$ |
| Population fraction of vaccinated people | $P_v$ | 0.8 | 0 to 1 |
| Vaccine efficacy | VE | 0.8 | 0 to 1 |
| Proportion of unvaccinated population with natural immunity | NI | 0.2 | 0 to 1 |
| Population of entire society | N | $10^7$ | $> 0$ |

**Table 2:** Quantities calculated from model results (mathematical definitions in Appendix 1, Section A1.3)

| Name | Symbol |
| --- | --- |
| Attack rate in the vaccinated population | $A_v$ |
| Attack rate in the unvaccinated population | $A_u$ |
| Attack rate in the overall population (vaccinated and unvaccinated) | $A_t$ |
| Share of infections among vaccinated people that were due to contacts with infectious unvaccinated people | $B_v$ |

### Analysis

The attack rate among the vaccinated population is defined as the proportion of initially-susceptible vaccinated people who become infected during the epidemic:

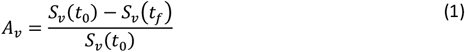

where *S*_*v*_(*t*_0_) is the number of susceptible vaccinated people at the beginning of the epidemic and *S*_*v*_(*t*_*f*_) is the number of susceptible vaccinated people remaining once there are no longer any infectious people in the entire (vaccinated and unvaccinated) population. *A*_*u*_ is defined equivalently, for the unvaccinated, replacing the *v* subscripts with *u* in Eq. 1. The overall attack rate for the full (vaccinated plus unvaccinated population) is:

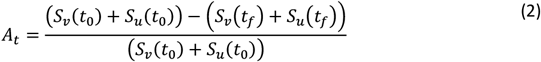

We also define *B*_*v*_ as the share of infections among vaccinated people that were due to contacts with infectious unvaccinated people (see Eq. A6 of Appendix 1).

We focus on segregation types that are targeted at the unvaccinated group. We assume, for simplicity, that segregation has no impact on the contact frequency of vaccinated people (*m*_*v*_ = 0). We also assume that the contact frequencies in both groups are the same when there is no segregation 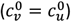. We use the same values as used by Fisman et al. [9] for the remaining parameters: *P*_*v*_ = 0.8, *VE*=0.8, *NI*=0.2, γ = 73 yr^-1^, and *N* = 10^7^. These values were intended to be representative for COVID-19 and vaccination; in particular, the recovery rate of 73 yr^-1^ is equivalent to a recovery time of 5 days [22,23], and assumed to be the same for vaccinated and unvaccinated people.

Appendix 2 contains supplementary figures with results for different parameter combinations, including *P*_*v*_ ≠ 0.8, *m*_*v*_ ≠ 0 and 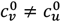. In all results in this paper, simulations were initiated with a seed number of 100 infectious individuals distributed proportionately among the two sub-populations.

## Results

Fig. 1 shows simulation results for a range of model parameters for different epidemiological conditions and degrees and types of societal segregation. Each row of panels is for a fixed value of 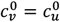. Moving from the top row (Figs. 1a.i to a.iv) to the bottom (Figs. 1e.i to e.iv), 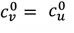. The left column of panels shows how the attack rate among the vaccinated population, *A*_*v*_, changes with the degree of segregation, η. The second and third columns show *A*_*u*_ and *A*_*t*_ as functions of η, respectively, and the right column shows how *B*_*v*_, the share of vaccinated infections that were due to contacts with unvaccinated people, varies with η.

**Figure 1:**
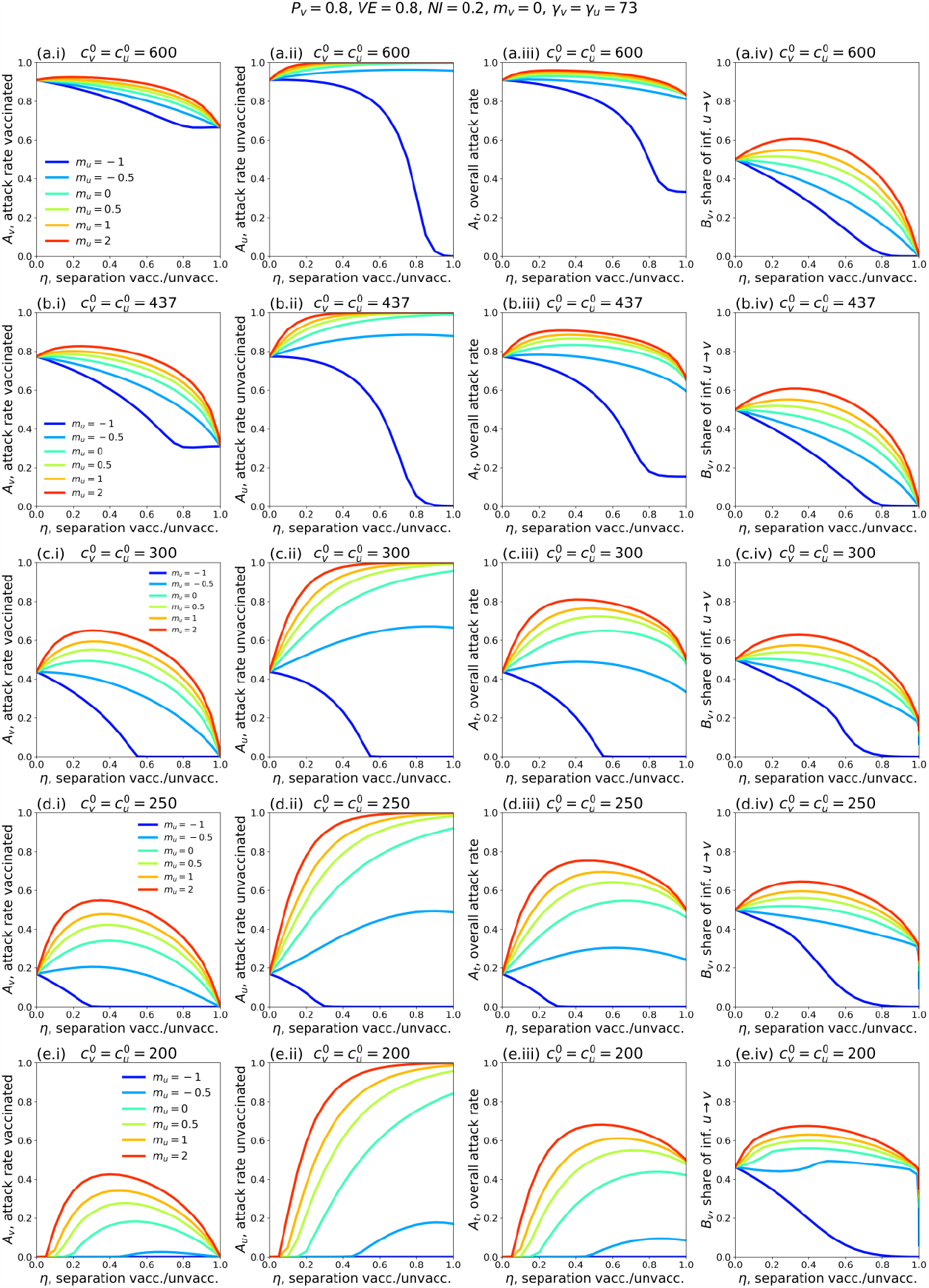
Attack rates *A*_*v*_ (vaccinated population), *A*_*U*_ (unvaccinated population), *A*_*t*_ (overall population) and share of vaccinated infections that were due to contacts with unvaccinated people, *B*_*v*_, as functions of the degree of segregation, η, of the vaccinated and unvaccinated. Each row of panels shows *A*_*v*_, *A*_*u*_, *A*_*t*_ and *B*_*v*_ for a particular choice of 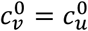. Values of fixed model parameters are indicated at the top of the figure. For reference, in a single-population (no vaccination) model, the corresponding *R*_*0*_ values for rows a-e of the figure are 8.2, 6.0, 4.1, 3.4 and 2.7, respectively.

Figs. 1c.i to c.iv show results for a moderate value of 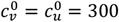 contacts/yr. For reference, in a single-population (no vaccination) model, *c* = 300 contacts/yr and γ_*v*_ = γ_*u*_ = 73 yr^-1^ corresponds to a basic reproduction number *R*_0_ = *c*/γ = 4.1.

In Fig. 1c.i, when *m*_*u*_ =-1 and *m*_*u*_ = -0.5 (reflecting large and moderate degrees of exclusion and isolation of unvaccinated people) the vaccinated attack rate, *A*_*v*_ decreases with increasing segregation. However, when *m*_*u*_ > 0 (compounding of unvaccinated people) or *m*_*u*_ = 0 (segregation has no influence on contact frequency of unvaccinated people), there is a maximum in *A*_*v*_ for moderate values of η. Therefore, with compounding segregation, large values of η are required for *A*_*v*_ to be lower than its value for no segregation (η = 0). Fig. 1c.ii shows that the unvaccinated attack rate, *A*_*u*_, increases with segregation for anything other than strong isolating segregation (*m*_*u*_ approaching -1). This produces a maximum in the overall attack rate *A*_*t*_ at moderate degrees of segregation, even for values of *m*_*u*_ for which *A*_*v*_ decreases monotonically (*m*_*u*_ = -0.5). Fig. 1c.iv shows that *B*_*v*_, the share of vaccinated infections that are due to unvaccinated people, has a shape similar to *A*_*v*_(η, *m*_*u*_). In all panels, 20% of the total population is unvaccinated (*P*_*v*_ = 0.8; Table 1).

Figs. 1c.i to c.iv therefore demonstrate that whether segregation increases or decreases the vaccinated-population attack rate depends on both the degree of segregation and how segregation affects contact frequency.

Figs. 1a.i to a.iv and 1b.i to b.iv show results for larger 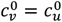. Compared to Fig. 1c.i, *A*_*v*_ in Figs. 1a.i and 1b.i does not increase much with η when *m*_*u*_ > 0, and *A*_*v*_ no longer has a maximum when *m*_*u*_ = 0. It can also be seen that *A*_*v*_ increases with increasing 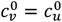 when there is no segregation (η = 0).

Reducing 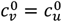 (Figs. 1d.i to d.iv and 1e.i to e.iv), decreases *A*_*v*_(η = 0), and larger η can dramatically increase *A*_*v*_. Even with an isolating segregation policy (*m*_*u*_ = −0.5 in Fig. 1d.i), *A*_*v*_ is increased for moderate values of η.

When 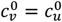 are small enough (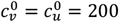 contacts/yr in Fig. 1e.i, corresponding to *R*_*0*_ = 2.7 in a single population (no vaccination) model), there is no epidemic among the vaccinated in the absence of segregation (*A*_*v*_(η = 0) = 0). However, a non-zero vaccinated-population attack rate (*A*_*v*_ > 0) occurs if η is sufficiently large, and emerges regardless of whether one isolates or compounds the unvaccinated. Therefore, for small enough values of 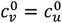, any segregation could increase infections among the vaccinated.

The main qualitative features of the above results for *P*_*v*_ = 0.8 hold for other values of *P*_*v*_. Appendix 2 provides a detailed exploration of results for *P*_*v*_ = 0.1 through 0.99; and for two values of VE (0.4 and 0.8). When VE is decreased, *A*_*v*_ is not strongly influenced by η, regardless of *m*_*u*_; therefore, any beneficial effect of segregation on *A*_*v*_ is reduced as VE decreases.

Appendix 2 also explores 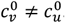. For example, when 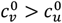, the unvaccinated contact frequency is reduced even when there is no segregation; increasing η can then increase *A*_*v*_ substantially compared to the case of 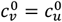, holding all other parameter values constant (see panels a.i and b.i in Figs. A2.28 and A2.31).

## Discussion

Segregation can have substantially different and negative impacts on the outcome of an epidemic, depending on the type and degree of segregation, and depending on cultural and population-density factors, for example, that co-determine 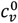 and 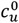.

Segregation that compounds the unvaccinated (*m*_*u*_ > 0 and *m*_*v*_ = 0) generally causes an increase in the vaccinated-population attack rate, *A*_*v*_, for small and intermediate degrees of segregation, η, while for large η, *A*_*v*_ decreases below its value in an unsegregated society. Segregation that isolates and excludes the unvaccinated (*m*_*u*_ < 0 and *m*_*v*_ = 0) decreases *A*_*v*_ for “more contagious viruses” (i.e. large 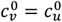, large *R*_*0*_); however, for “less contagious viruses” (smaller 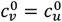, smaller *R*_*0*_), both isolating and compounding types of segregation can increase *A*_*v*_ beyond its value in an unsegregated society. For “viruses that are not very contagious” (small 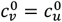, small *R*_*0*_), applying segregation can cause a sizeable epidemic among the vaccinated even though virtually no vaccinated people would be infected in an unsegregated society. Segregation increases the unvaccinated attack rate, *A*_*u*_, for compounding and moderately isolating types of segregation, and *A*_*u*_ is only decreased for strongly isolating segregation (*m*_*u*_ approaching -1).

Except for large negative values of *m*_*u*_, and small unvaccinated population fractions, applying segregation has the effect of increasing the frequency of unvaccinated-to-unvaccinated contacts (see Appendix 1, Fig. A1.3). This increases the overall probability of a susceptible-infectious interaction, since the unvaccinated population has a higher fraction of susceptibles, and creates a form of core group dynamics [24-26]. At the same time, increasing segregation shields the vaccinated population from the increased prevalence of infection in the unvaccinated population. This trade-off causes the non-monotonic relationship between *A*_*v*_ and η. The same dynamic causes the emergence of an epidemic for large η when 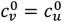 (and thus *R*_*0*_) is small.

We find that *B*_*v*_, the share of vaccinated infections that are due to contact with unvaccinated people, follows a similar trend to *A*_*v*_ as a function of the degree of segregation, when segregation has no impact on the vaccinated contact frequency (*m*_*v*_ = 0). For this type of segregation, *A*_*v*_ and *B*_*v*_ either increase or decrease simultaneously with increasing η, depending on the value of *m*_*u*_, and *B*_*v*_ is minimized for complete segregation. When *m*_*v*_ = 0, there is no type or degree of segregation that reduces the vaccinated attack rate while simultaneously increasing the risk to vaccinated people from unvaccinated people (Fig. 1). Therefore, there are no circumstances in which the unvaccinated cause a disproportionate risk to the vaccinated, contrary to conclusions in Fisman et al. [9].

In contrast, when *m*_*v*_ ≠ 0, such that segregation affects the contact frequencies of vaccinated people, increasing segregation can cause *A*_*v*_ to increase while *B*_*v*_ decreases and vice-versa (see Appendix 2, Figs. A2.25 and A2.26).

The impact of vaccination-status-based societal segregation on contact frequencies has not previously been considered to our knowledge, including in network-based models in which unvaccinated people cluster together in “cliques” or households [26-28].

### Limitations

Our model assumes only two risk populations (vaccinated and unvaccinated), considers only the attack rates on epidemic completion (*A*_*v*_, *A*_*u*_ and *A*_*t*_), and takes the degree of segregation η to be time-independent, without variation due to public holidays and such. It does not consider other outcomes such as death or hospitalization. Our model assumes an all-or-nothing VE, without waning immunity or influence on infectiousness; and no possibility of reinfection. We do not consider the impact of segregation policies on vaccination rates. SIR models and their variations are based on the paradigm of transmission due to pairwise contact between a recently infected and a susceptible individual. However, this paradigm is unable to account for important features of viral respiratory disease incidence, in particular its rapid emergence and disappearance occurring at essentially the same time at widely dispersed locations [29]. Air-borne transmission via suspended aerosol particles is not directly compatible with pairwise transmission, since it occurs in built environments where many people may transit or be present [30]. A related and unavoidable limitation is the lack of reliable empirical evaluations of needed infectious contact frequencies, which is important because our calculated outcomes are sensitive to the chosen contact frequency values. Lastly, we do not consider the deleterious health impacts of the segregation policies themselves, which can be significant [32-38].

## Conclusion

In the two-population mixing-model framework, vaccination-status-based societal segregation can lead to substantially different and counter-intuitive epidemic outcomes depending on the type and degree of segregation, and depending on complex cultural and physical factors that co-determine infectious contact frequencies (i.e., the products βc). Negative epidemiological consequences can occur for either segregated group, irrespective of the deleterious health impacts of the policies themselves.

Given the lack of reliable empirical evaluations of needed infectious contact frequency values, given the demonstrated outcome sensitivities to the infectious contact frequencies, and given the intrinsic limitations of SIR models in this application, we cannot recommend that SIR modelling be used to motivate or justify segregation policies regarding viral respiratory diseases, in the present state of knowledge.

## Data Availability

This is a modeling paper, therefore all data is from simulations. Information for reproducing the simulations is contained in the manuscript and appendices.

### Appendix 1: Elaboration of the model

This is Appendix 1 of the pre-print *“*Viral respiratory epidemic modeling of societal segregation based on vaccination status” by J. Hickey & D.G. Rancourt, uploaded to https://www.medrxiv.org/ on 2023-10-31 (Version 5).

#### A1.1: Model differential equations and “mixing” rule

The model is a susceptible-infectious-recovered (SIR) model with two populations: vaccinated (subscript *v*) and unvaccinated (subscript *u*) people, consisting of the following six differential equations:

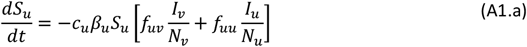

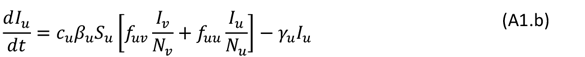

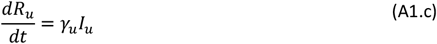

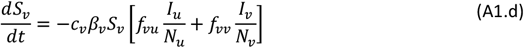

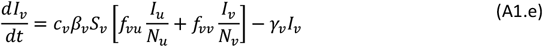

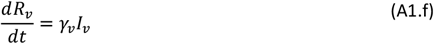

*S*_*u*_, *I*_*u*_, and *R*_*u*_ represent the number of susceptible, infectious, and recovered unvaccinated people, at time *t. N*_*u*_ represents the total number of unvaccinated people. *c*_*u*_ represents the contact frequency (number of contacts per unit time) of unvaccinated people. β_*u*_ is the probability that a susceptible unvaccinated person becomes infected upon contact with an infectious person (regardless of whether the infectious person is vaccinated or unvaccinated). γ_*u*_ is the rate at which infected unvaccinated people recover from infection. The quantities *S*_*v*_, *I*_*v*_, *R*_*v*_, *N*_*v*_, *c*_*v*_, β_*v*_, and γ_*v*_ are defined equivalently, for vaccinated people.

There are thus two “β parameters”, two “*c* parameters” and two “γ parameters” in our model. Since each β parameter always occurs as part of a product with its respective *c* parameter, the β parameters can freely be set equal to 1: this imposes that the “contacts” considered in the model are, by definition, only those contacts that are of sufficiently close proximity and long duration that an infection is guaranteed to occur when a susceptible and an infectious person meet. We set β_*v*_ = β_*u*_ = 1 in the main text, without any loss of generality.

*f*_*ij*_ is the probability that a person of type *i* (either *u* or *v*) has a contact with a person of type *j* (either *u* or *v*), and is defined as follows:,

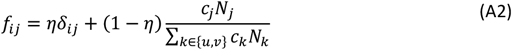

where 0 ≤ η ≤ 1 and δ_*ij*_ is the Kronecker delta, such that δ_*uu*_ = δ_*vv*_ = 1 and δ_*uv*_ = δ_*vu*_ = 0. η is therefore a parameter that controls the degree of separation between the *u* and *v* sub-populations. For example, when η = 1, then *f*_*uu*_ = *f*_*vv*_ = 1 and *f*_*uv*_ = *f*_*vu*_ = 0, such that *u* people only ever have contacts with other *u* people and likewise for *v* people (complete separation). At the other extreme, when η = 0, then the probability that a *u* person has a contact with a *v* person is entirely determined by the relative proportions of *u* and *v* people, weighted by their respective contact frequencies (no separation, or “random mixing”).

#### A1.2: Variation of contact frequency with degree of separation

To allow contact frequency to vary with degree of separation, we define:

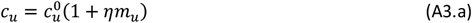

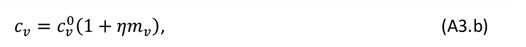

where 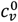 and 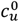 represent the contact frequencies for unvaccinated and vaccinated people when there is no separation (η = 0), and *m*_*u*_ ≥ −1 and *m*_*v*_ ≥ −1 are two parameters that control how non-zero separation impacts the contact frequencies of unvaccinated and vaccinated people.

Fig. A1.1 shows *c*_*u*_ as a function of η, for different values of the parameter *m*_*u*_ and for 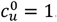. As can be seen, when 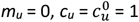, such that the contact frequency is constant regardless of the degree of separation.

**Fig. A1.1:**
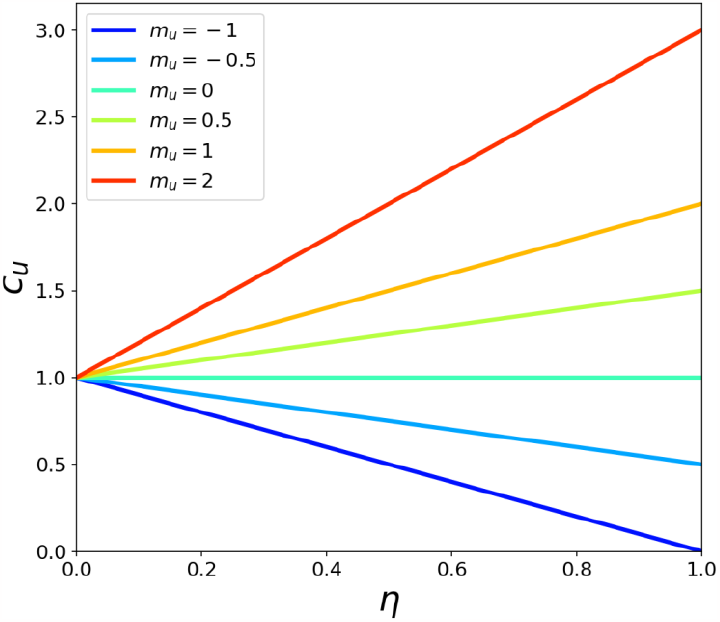
Variation of the unvaccinated contact frequency, *c*_*u*_, with degree of separation, η, for various values of the parameter *m*_*u*_. 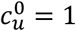 in the figure.

When *m*_*u*_ < 0, the contact frequency of unvaccinated people decreases with increasing η. This represents a separation policy that excludes unvaccinated people from public spaces while also isolating them from themselves to some degree. For example, a pair of intermediate values of *m*_*u*_ < 0 and 0 < η < 1 could represent a separation policy (such as with “vaccination passports”) that excludes unvaccinated people from recreational venues like restaurants and cinemas but not more essential services such as grocery stores and hospitals. On the other hand, in the extreme case of *m*_*u*_ = −1 and η = 1, *c*_*u*_ = 0 such that unvaccinated people are completely separated and isolated, having no contacts with anyone.

When *m*_*u*_ > 0, the contact frequency of unvaccinated people increases with increasing η. This represents a separation policy that compounds unvaccinated people by placing them together in close quarters, for example in designated facilities for returning unvaccinated travelers.

Via the two parameters *m*_*u*_ and η, the model therefore spans the full range of contact frequencies, from *c*_*u*_ = 0 in the isolating extreme of *m*_*u*_ = −1 and η = 1 to arbitrarily high contact frequency in the compounding extreme with η = 1 and a *m*_*u*_ > 0.

The impact of separation on the vaccinated sub-population is independently controlled via the parameters 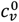 and *m*_*v*_.

#### A1.3 Intra- and inter-population contact frequencies

The contact frequency *c*_*i*_ represents the frequency of contacts for a person in group *i* (either *u* or *v*) irrespective of whether the contact partners belong to group *u* or *v*. We use the term “population-specific contact frequency” to refer to *c*_*i*_.

Given the probability *f*_*ij*_ that a person of type *i* has a contact with a person of type *j* (see Eq. A2), one can define the intra-population contact frequencies *c*_*ii*_ and the inter-population contact frequencies *c*_*ij*_ as follows:

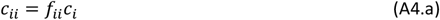

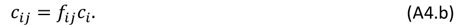

Eqs. A4 also show that *c*_*i*_ = *c*_*ii*_ + *c*_*ij*_.

Fig. A1.2 shows how the population-specific contact frequencies *c*_*u*_ and *c*_*v*_, the intra-population contact frequencies *c*_*uu*_ and *c*_*vv*_, and the inter-population contact frequencies *c*_*uv*_ and *c*_*vu*_ vary with η, for the six values of *m*_*u*_ explored in the main text and *m*_*v*_ = 0, and for three values of the population fraction of vaccinated people, *P*_*v*_.

Fig. 1 of the main text uses a value of *P*_*v*_ = 0.8, corresponding to the right column of panels in Fig. A1.2. Model results for *P*_*v*_ = 0.25 (left column of Fig. A1.2) and *P*_*v*_ = 0.5 (middle column of Fig. A1.2) are included in Appendix 2.

**Fig. A1.2:**
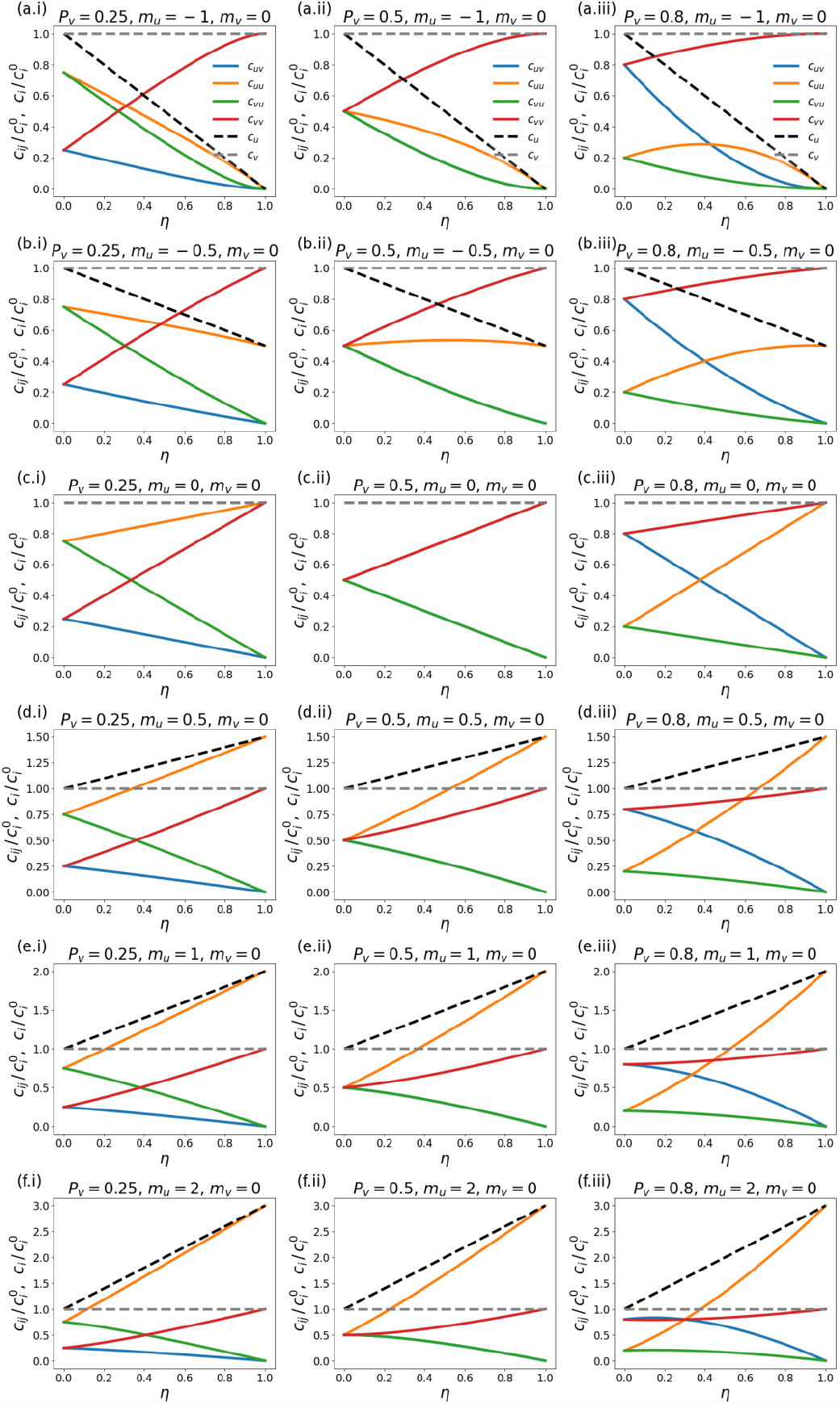
Normalized population-specific contact frequencies *c*_*u*_ and *c*_*v*_, intra-population contact frequencies *c*_*uu*_ and *c*_*vv*_, and inter-population contact frequencies *c*_*uv*_ and *c*_*vu*_, as functions of η, for the six values of *m*_*u*_ explored in the main text (each row of panels corresponds to a different value of *m*_*u*_), for *m*_*v*_ = 0, and for *P*_*v*_ = 0.2 (left column of panels), *P*_*v*_ = 0.5 (middle column), and *P*_*v*_ = 0.8 (right column).

#### A1.4: Quantities calculated from simulation results

The attack rate among the vaccinated population, *A*_*v*_, is defined as the proportion of initially-susceptible vaccinated people who become infected during the epidemic (Eq. 1 of the main text, reproduced below):

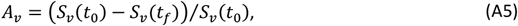

where *S*_*v*_(*t*_0_) is the number of susceptible vaccinated people at the beginning of the epidemic and *S*_*v*_(*t*_*f*_) is the number of susceptible vaccinated people remaining once there are no longer any infectious people in the entire (vaccinated and unvaccinated) population. *A*_*u*_ is defined equivalently, for the unvaccinated.

The quantity *B*_*v*_ is equal to the share of infections among vaccinated people that were due to contacts with infectious unvaccinated people, i.e.:

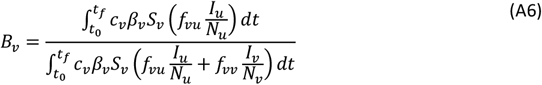

Similarly, *B*_*u*_ represents the share of infections among unvaccinated people that were due to contacts with infectious vaccinated people, and is defined in the same way as *B*_*v*_, (interchanging the *v* and *u* subscripts in Eq. A6).

### Appendix 2: Supplementary figures

This is Appendix 2 of the pre-print “Viral respiratory epidemic modeling of societal segregation based on vaccination status” by J. Hickey & D.G. Rancourt, uploaded to https://www.medrxiv.org/ on 2023-10-31 (Version 5).

#### A2.1: Epidemic curves

**Fig. A2.1:**
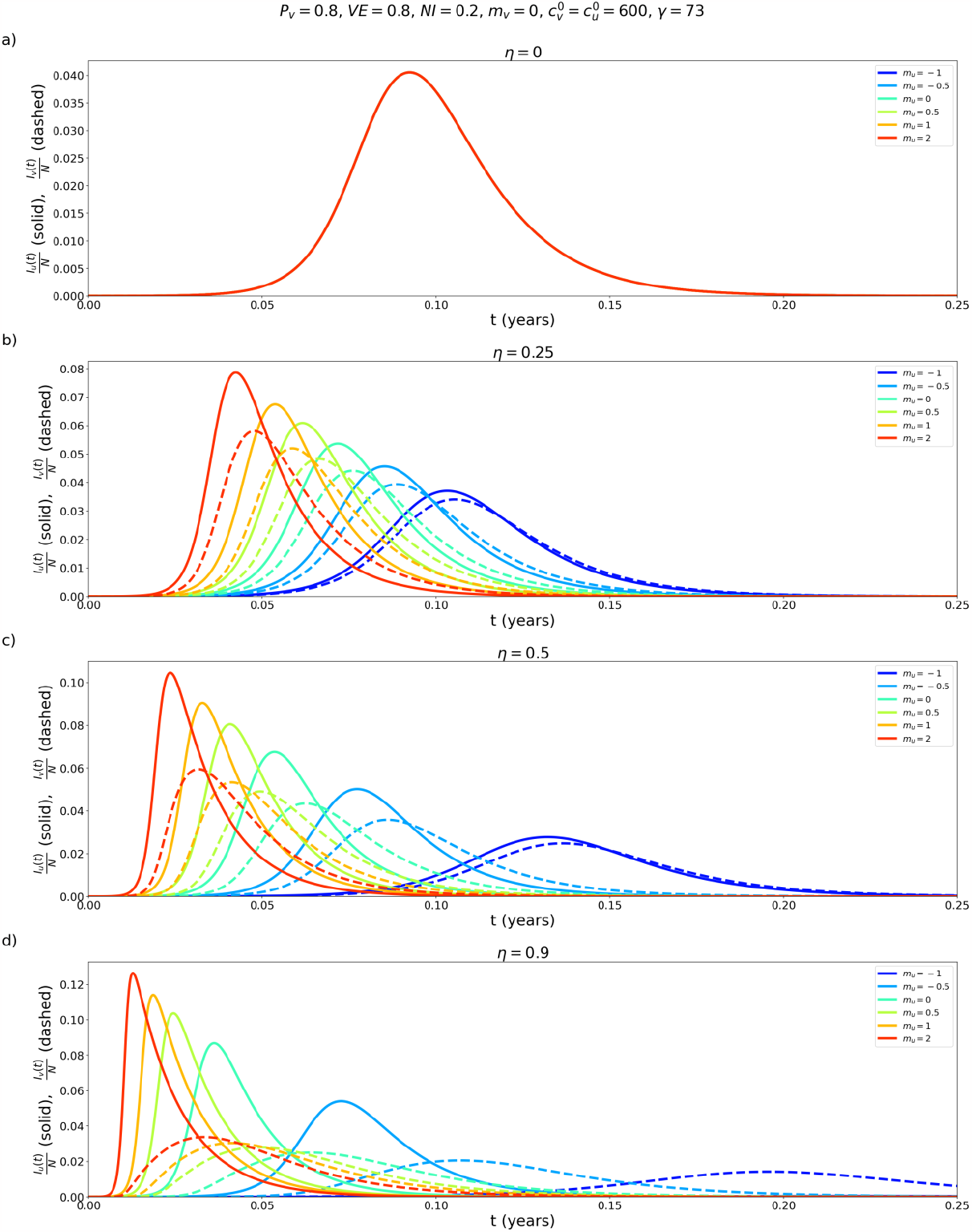
Population fraction of infectious individuals as a function of time, for the vaccinated (solid lines) and unvaccinated (dashed lines) populations, for parameters *P*_*v*_ = 0.8, VE = 0.2, NI = 0.2, m_v_ = 0, 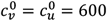, γ = 73. Each panel (a-d) shows a different value of η, and each coloured line is for a different value of *m*_*u*_, as indicated in the legend.

**Fig. A2.2:**
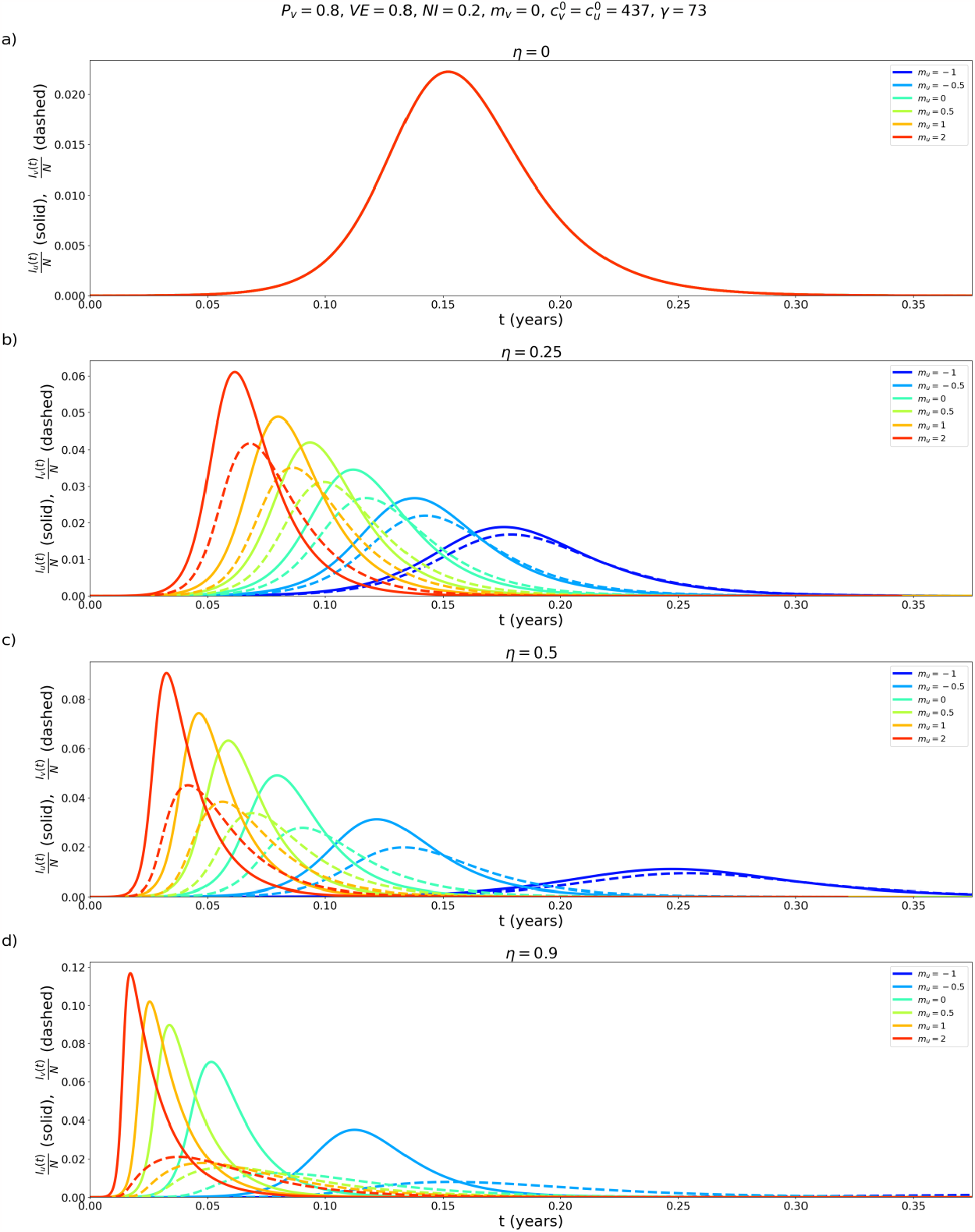
Same as Fig. A2.1, except that 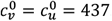.

**Fig. A2.3:**
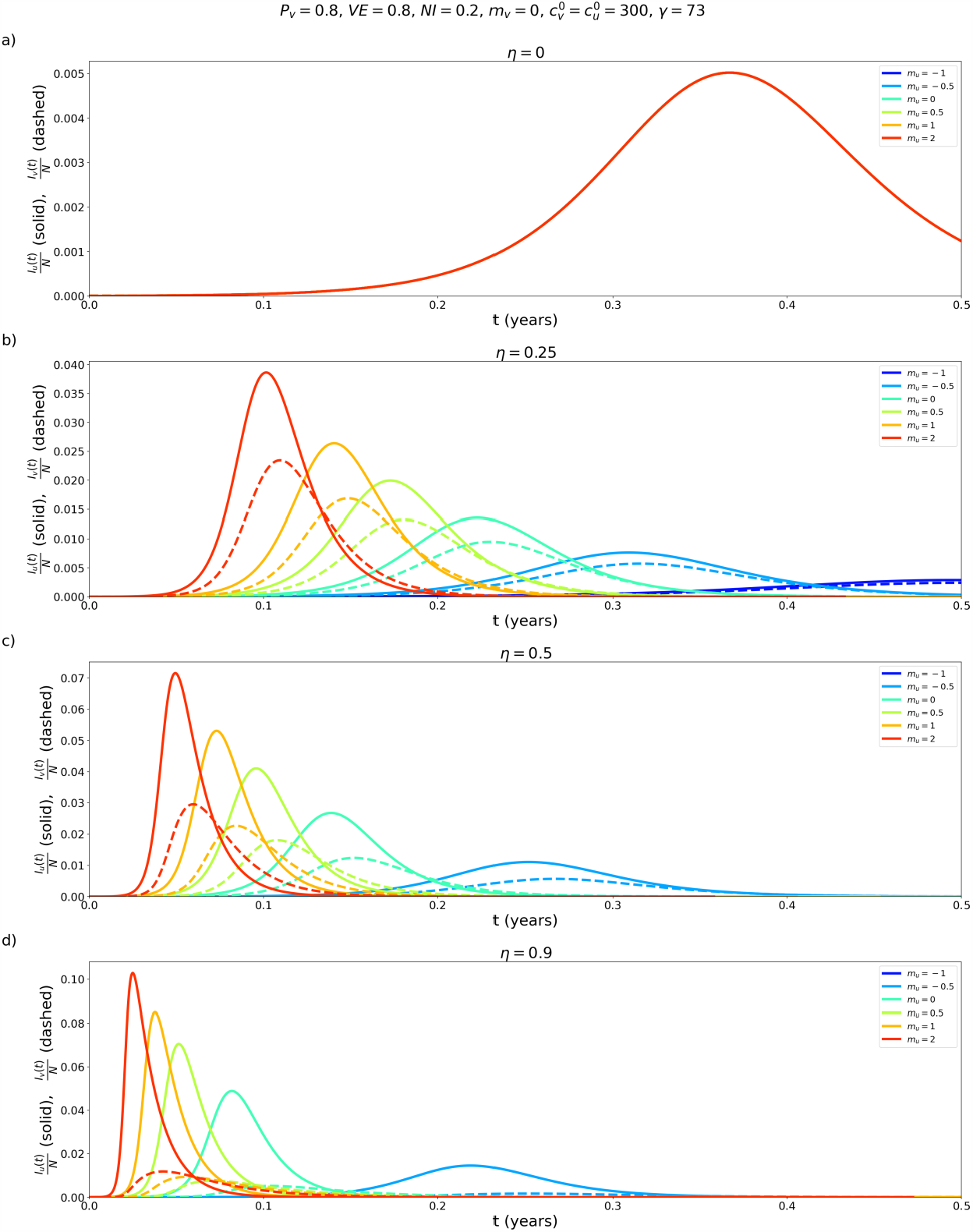
Same as Fig. A2.1, except that 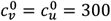.

**Fig. A2.4:**
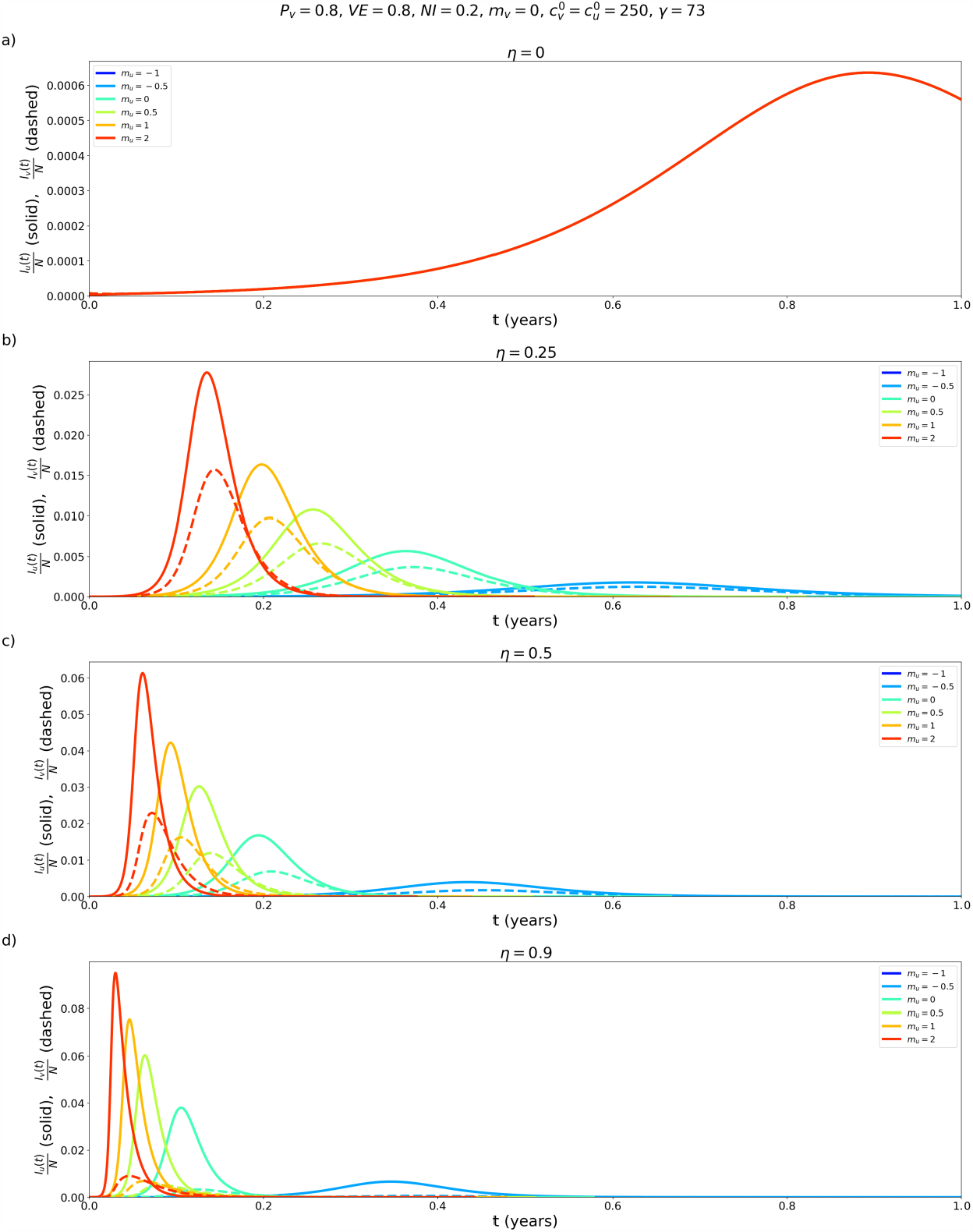
Same as Fig. A2.1, except that 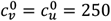.

**Fig. A2.5:**
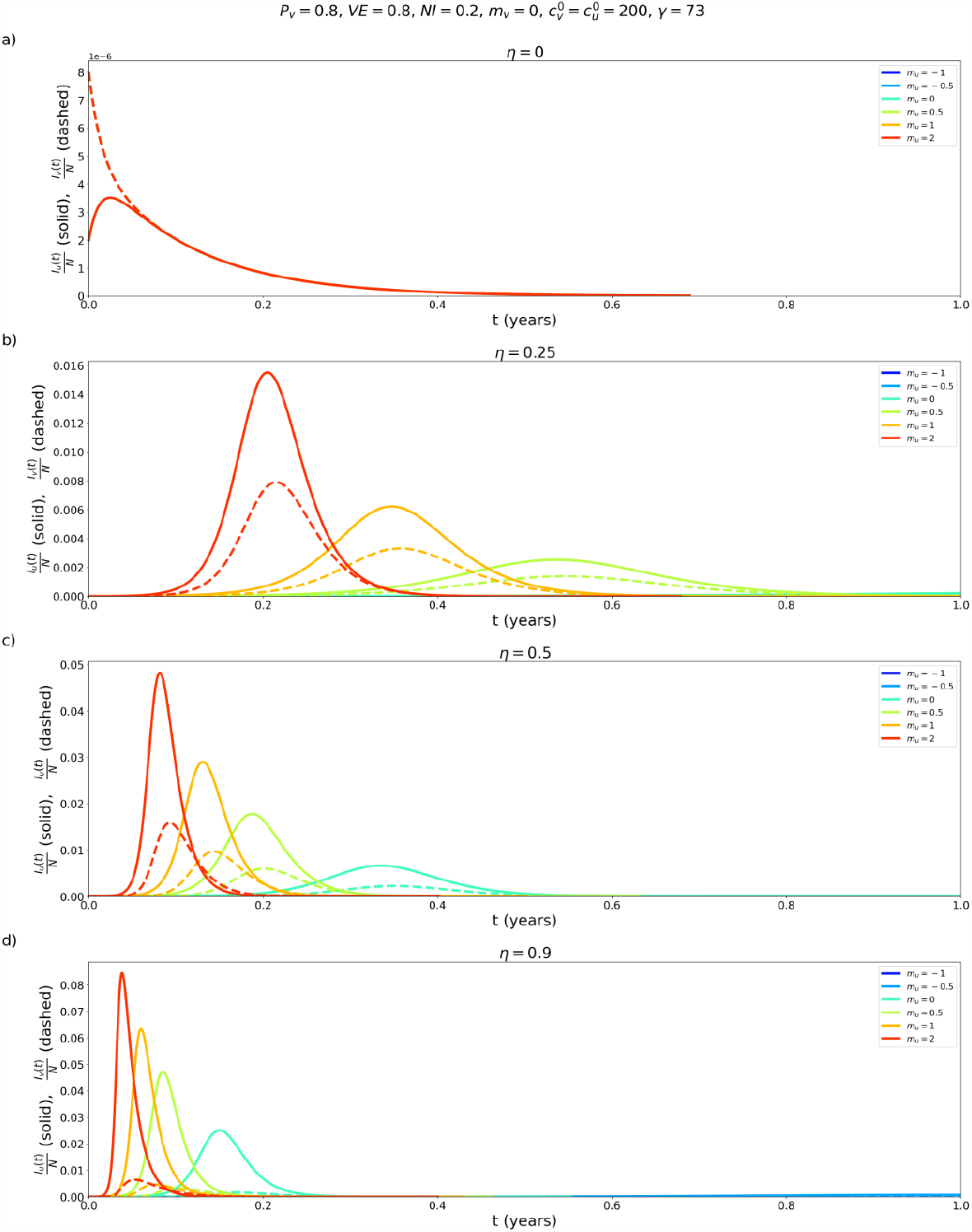
Same as Fig. A2.1, except that 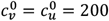.

#### A2.2: Epidemic outcomes for different values of *Pv*

##### A2.2.1: VE = 0.8

**Fig. A2.6:**
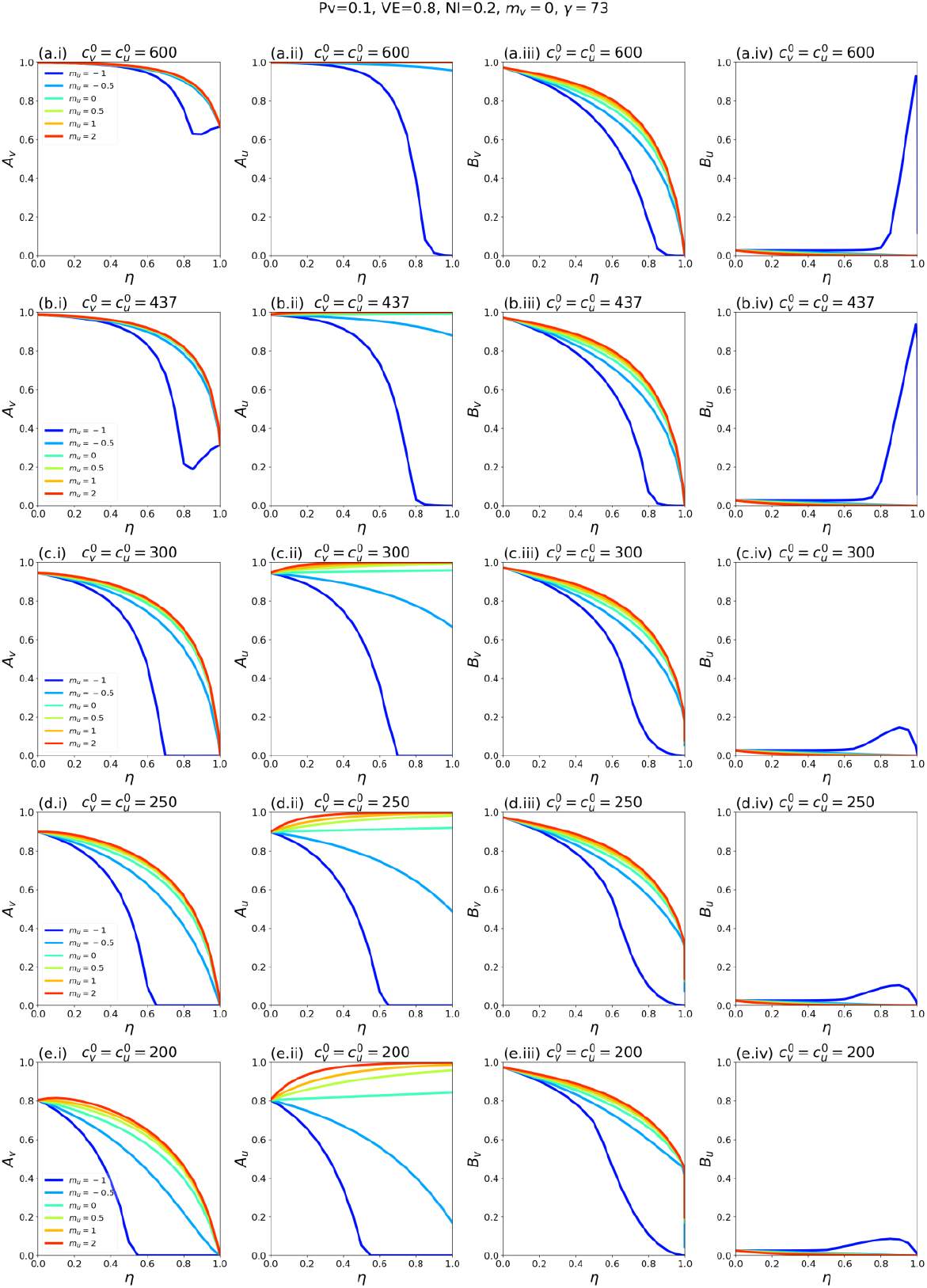
*A*_*v*_, *A*_*u*_, *B*_*v*_, and *B*_*u*_ as functions of η. Each row of panels corresponds to a choice of 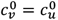, and each coloured line to a choice of *m*_*u*_ as indicated in the legends. For reference, in a single-population (no vaccination) model, the corresponding *R*_*0*_ values for rows a-e of the figure are 8.2, 6.0, 4.1, 3.4 and 2.7, respectively. Parameter values *P*_*v*_ = 0.1, VE = 0.8, NI = 0.2, *m*_*v*_ = 0, γ = 73.

**Fig. A2.7:**
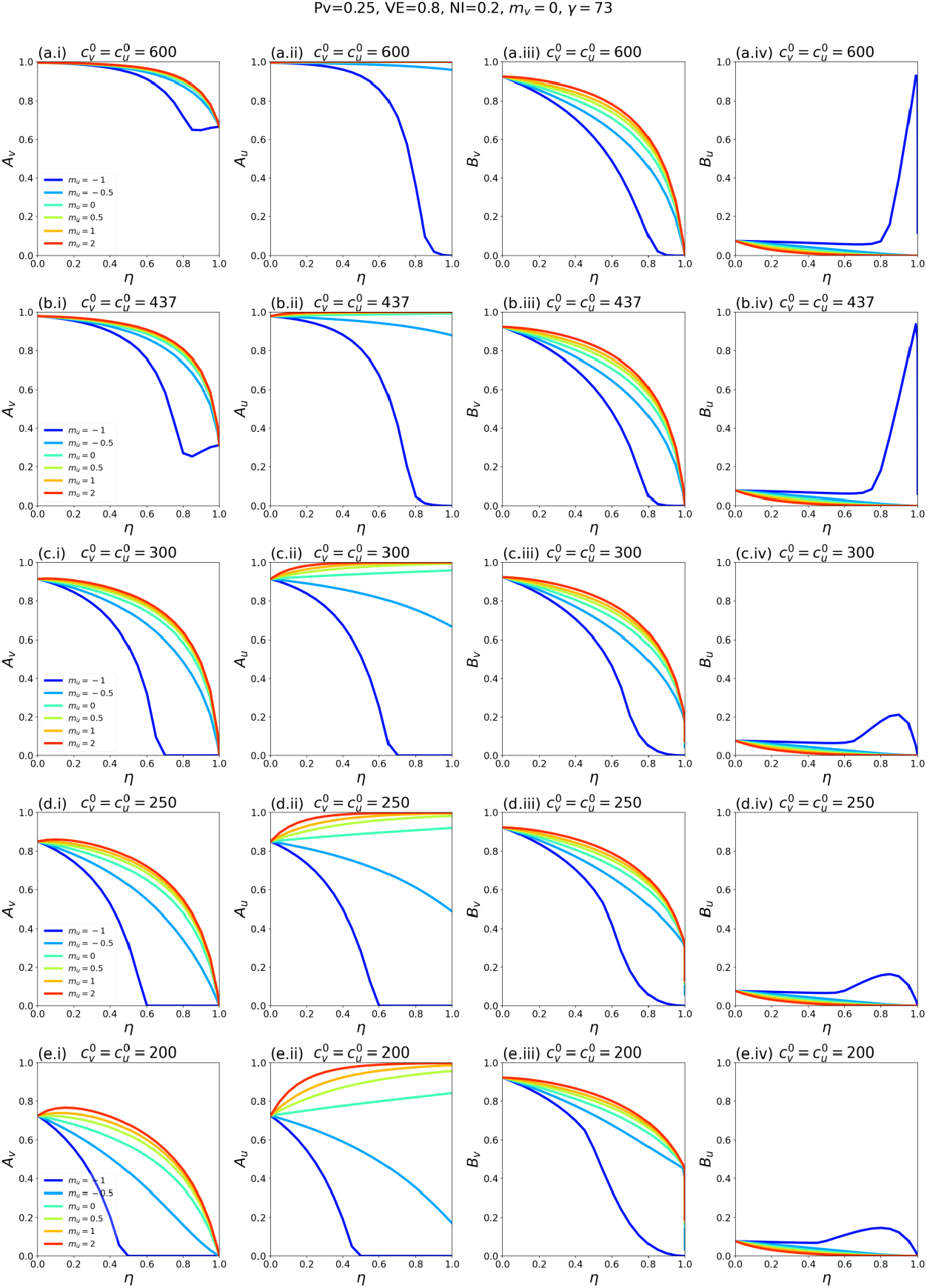
Same as Fig. A2.6, except that *P*_*v*_ = 0.25.

**Fig. A2.8:**
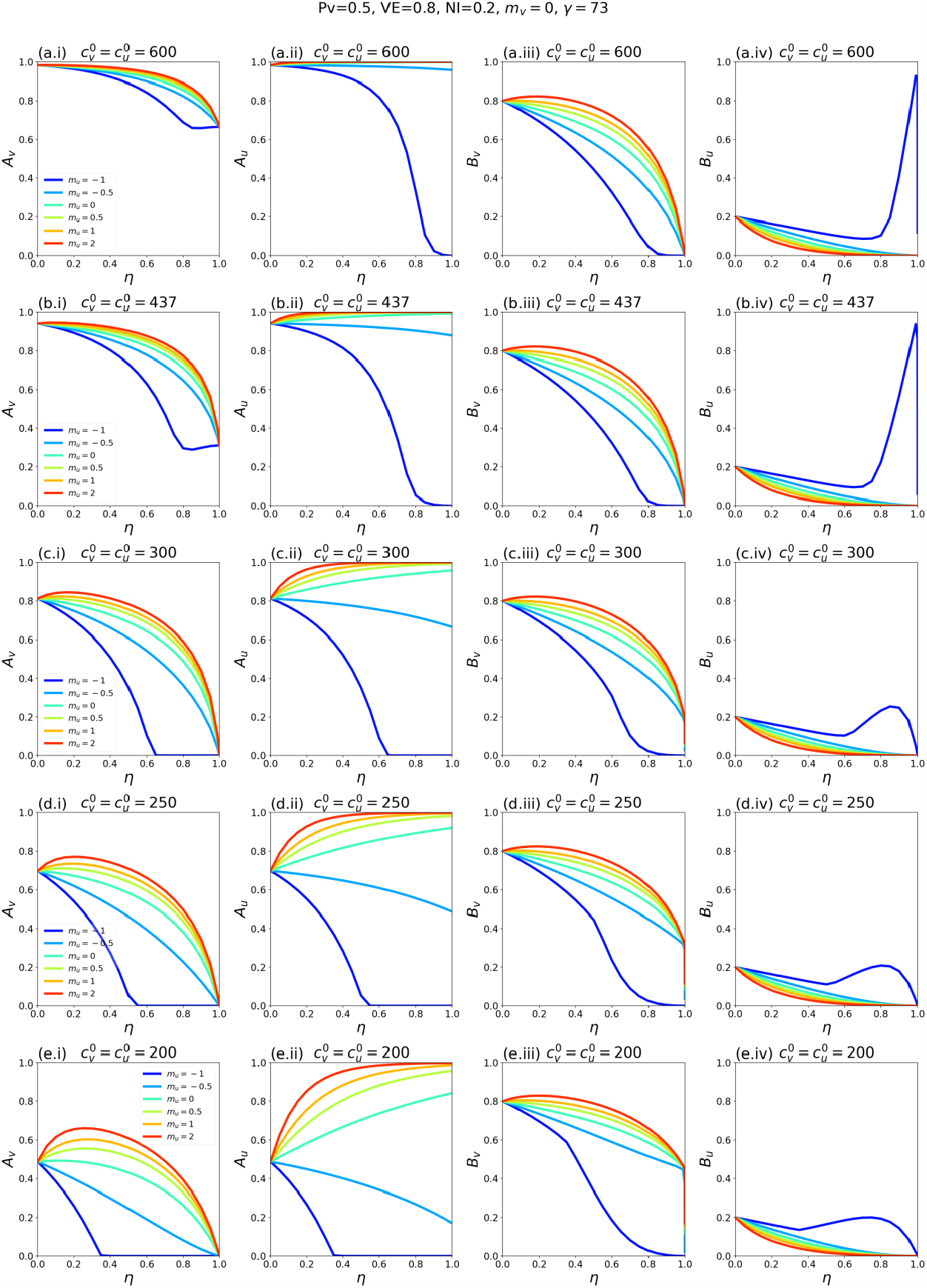
Same as Fig. A2.6, except that *P*_*v*_ = 0.5.

**Fig. A2.9:**
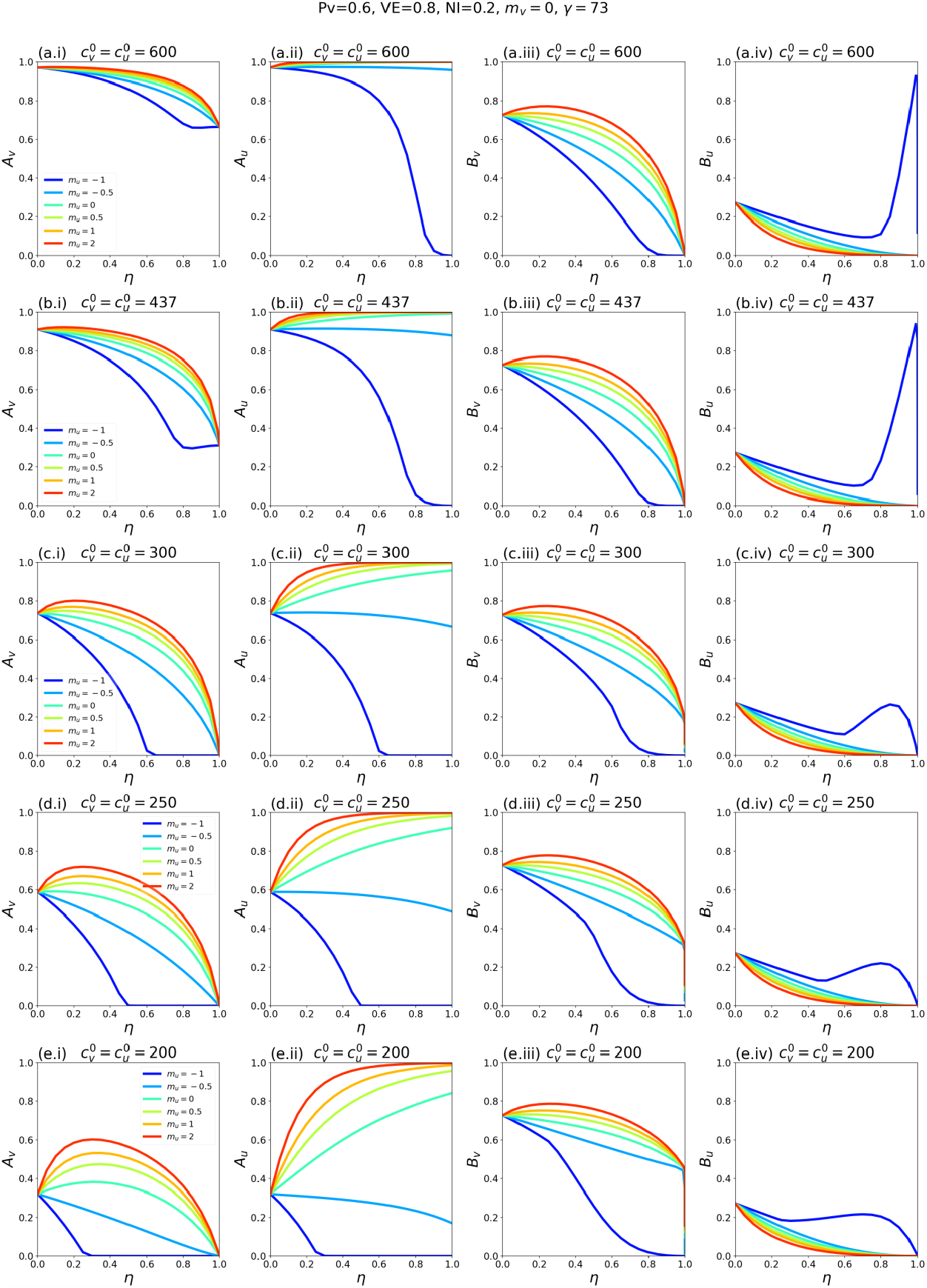
Same as Fig. A2.6, except that *P*_*v*_ = 0.6.

**Fig. A2.10:**
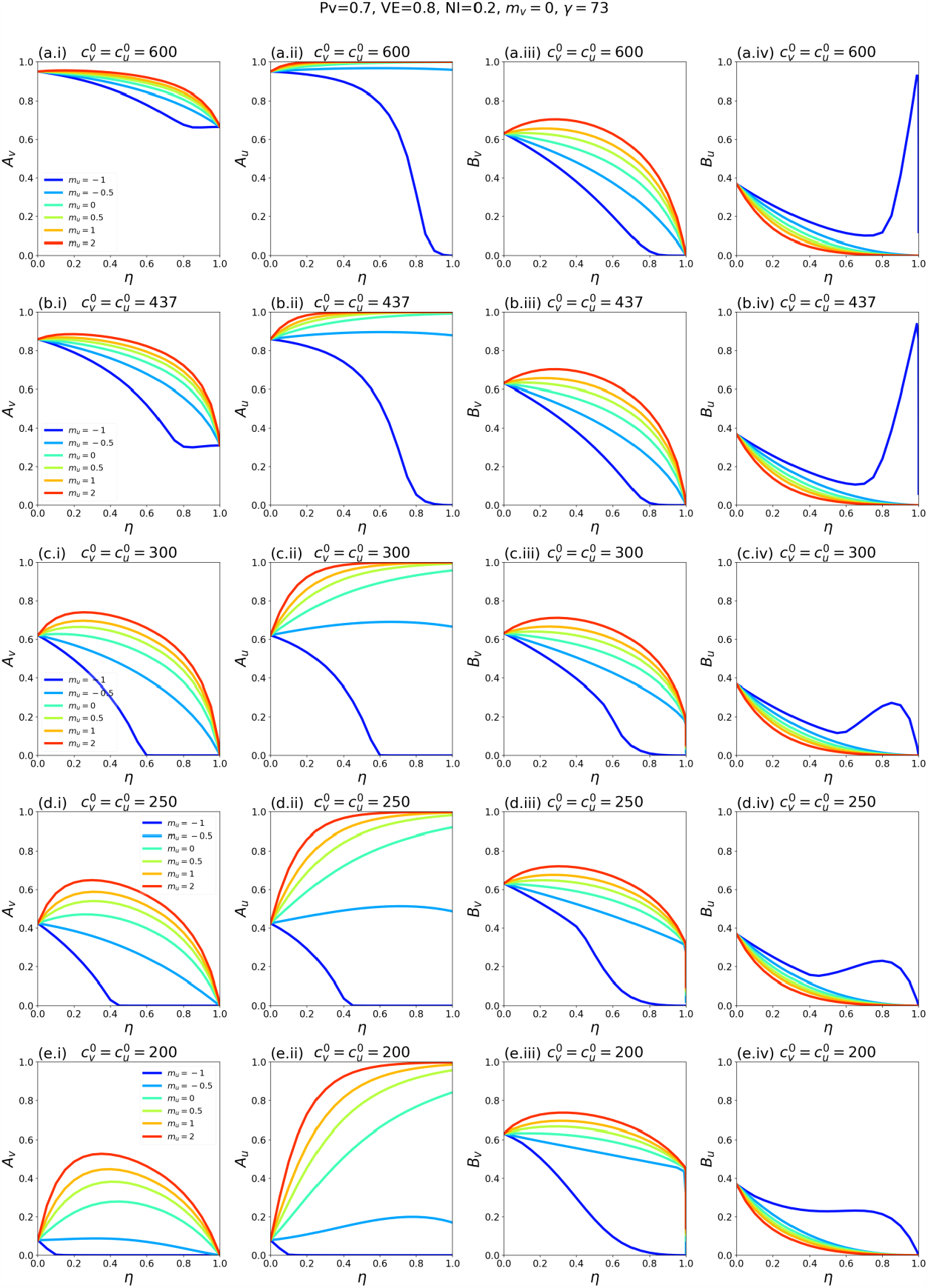
Same as Fig. A2.6, except that *P*_*v*_ = 0.7.

**Fig. A2.11:**
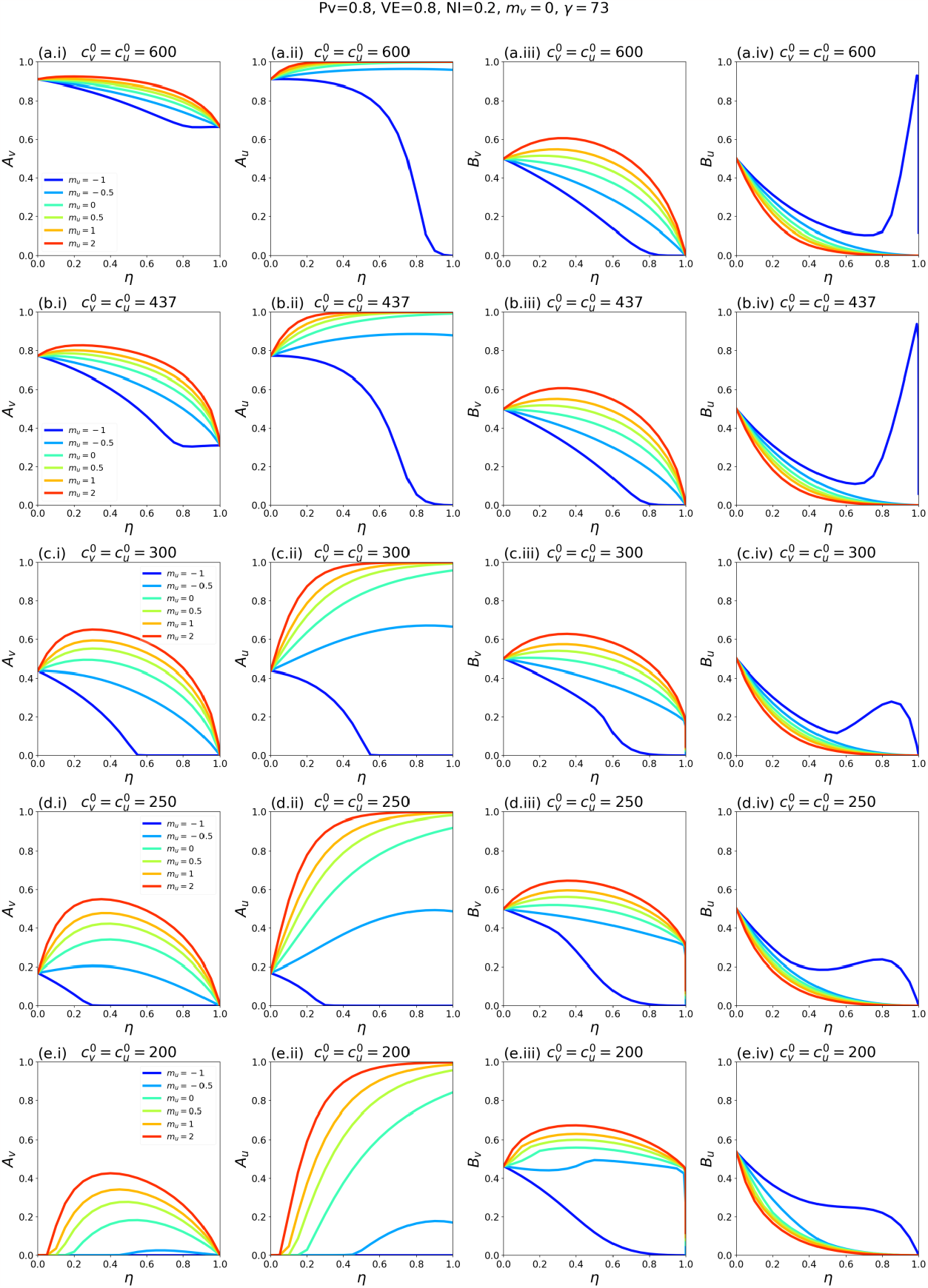
Same as Fig. A2.6, except that *P*_*v*_ = 0.8. This figure is for the same parameters as Fig. 1 of the main text, such that the *A*_*v*_, *A*_*u*_, and *B*_*v*_ columns in this figure are reproductions of the same columns in Fig. 1 of the main text.

**Fig. A2.12:**
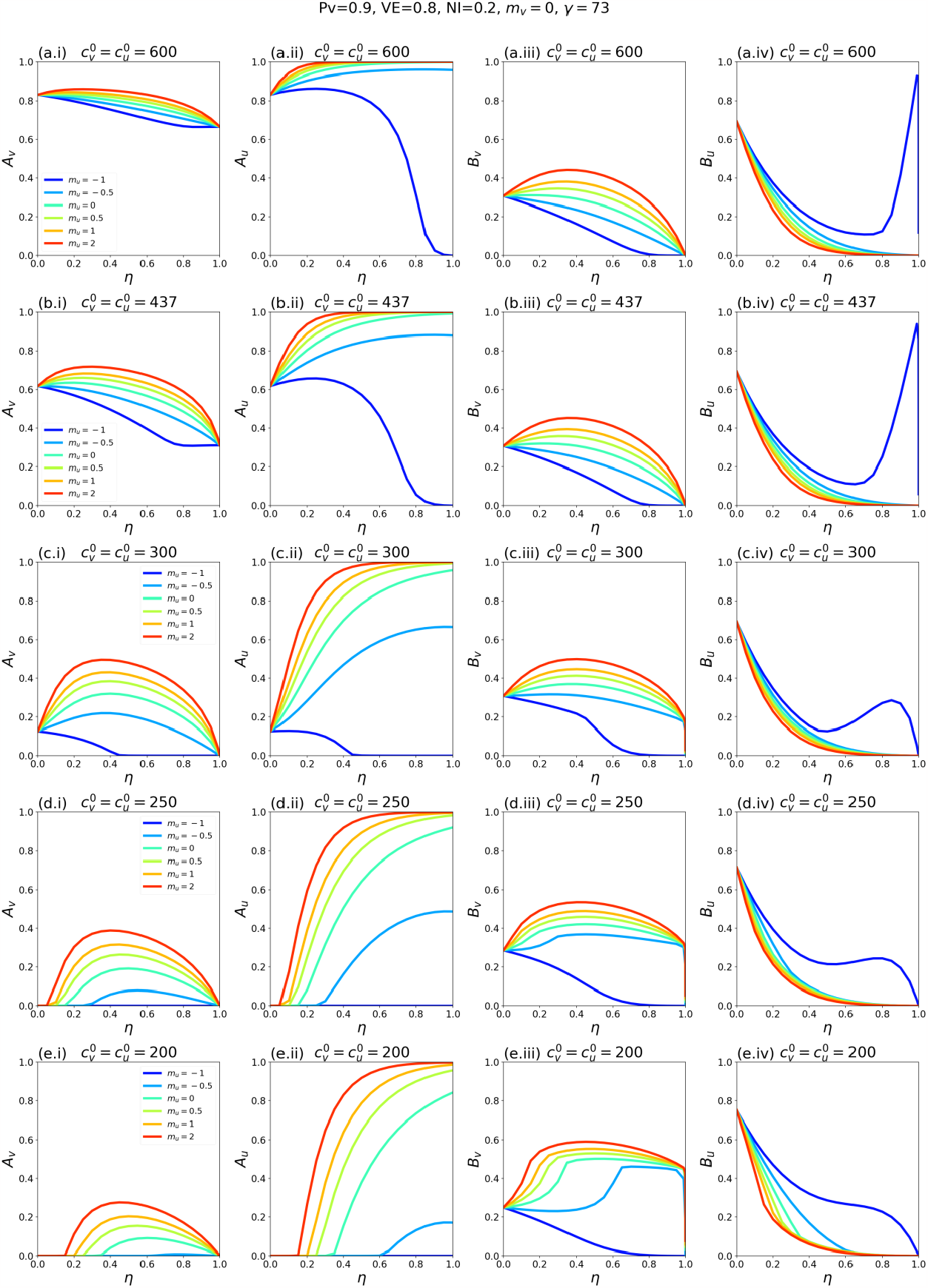
Same as Fig. A2.6, except that *P*_*v*_ = 0.9.

**Fig. A2.13:**
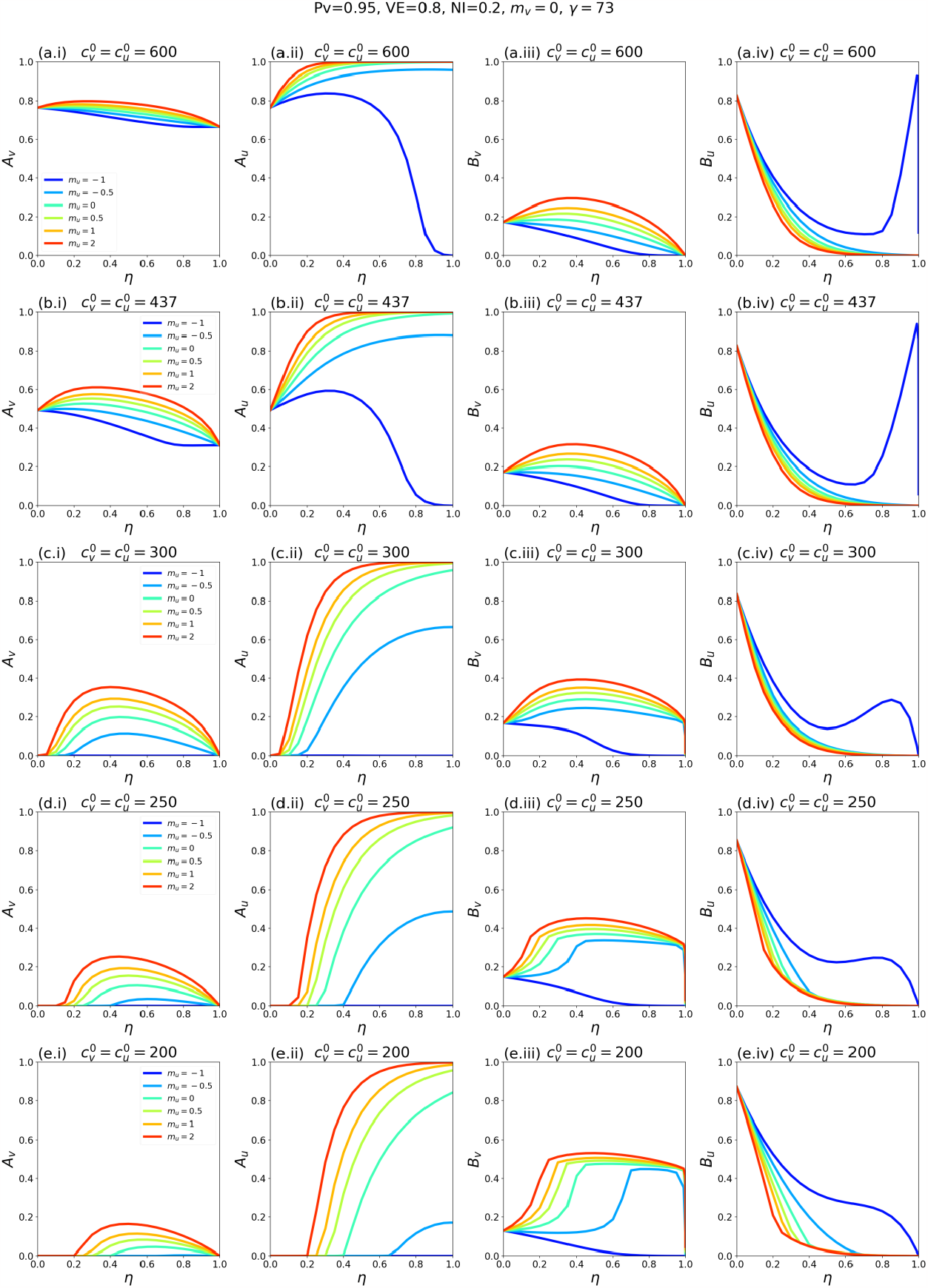
Same as Fig. A2.6, except that *P*_*v*_ = 0.95.

**Fig. A2.14:**
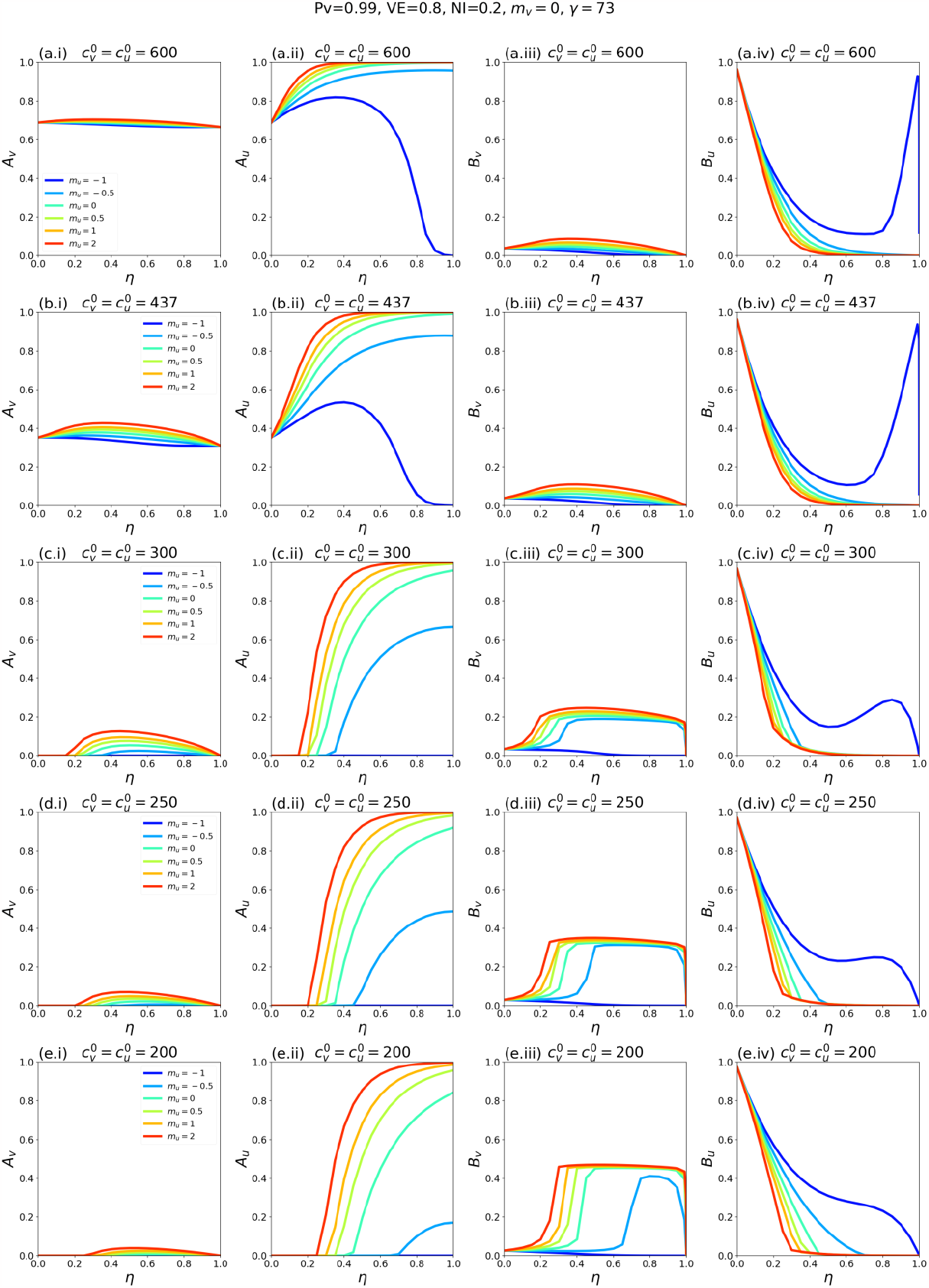
Same as Fig. A2.6, except that *P*_*v*_ = 0.99.

##### A2.2.2: VE = 0.4

**Fig. A2.15:**
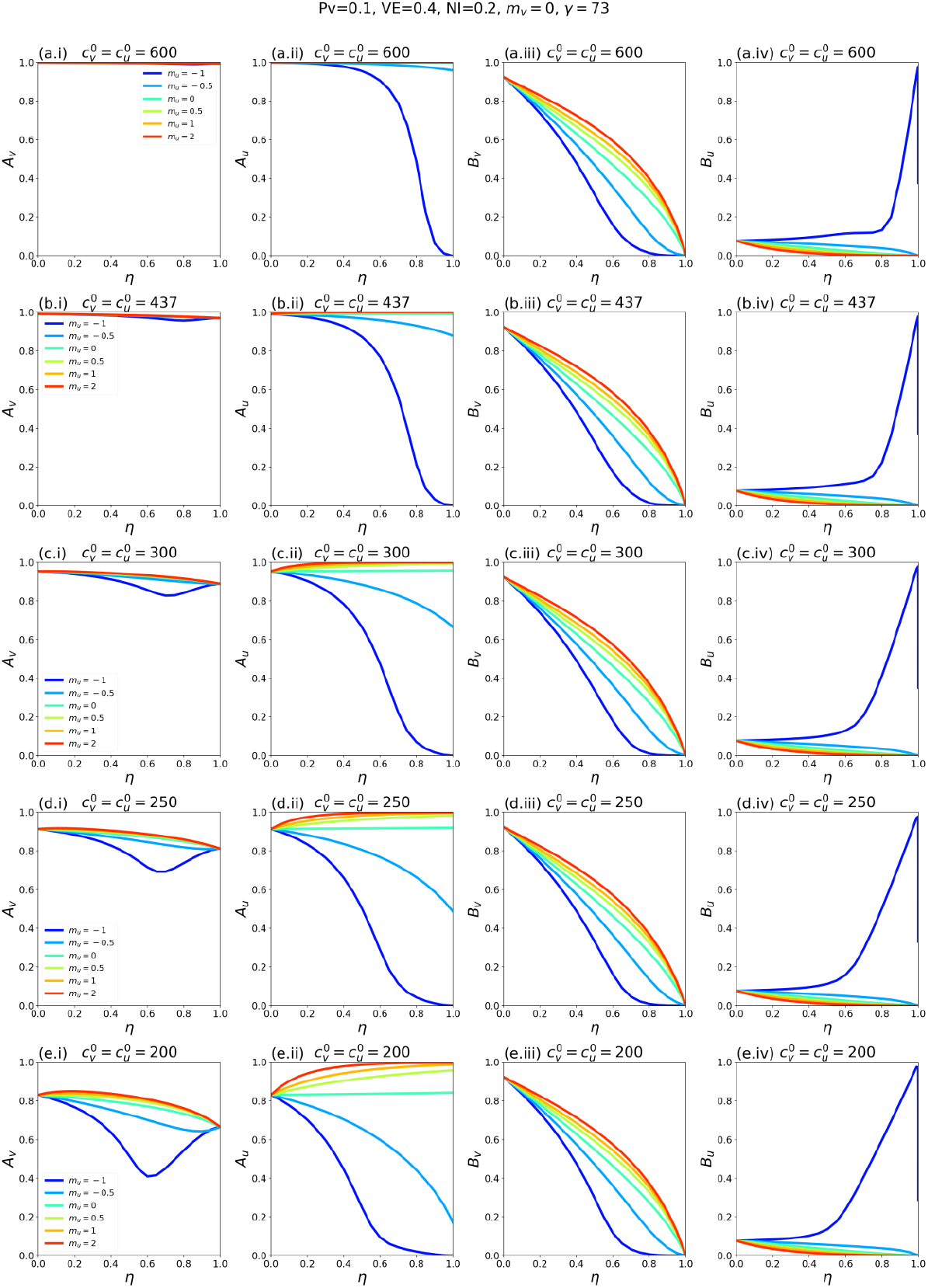
*A*_*v*_, *A*_*u*_, *B*_*v*_, and *B*_*u*_ as functions of η. Each row of panels corresponds to a choice of 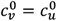, and each coloured line to a choice of *m*_*u*_ as indicated in the legends. For reference, in a single-population (no vaccination) model, the corresponding *R*_*0*_ values for rows a-e of the figure are 8.2, 6.0, 4.1, 3.4 and 2.7, respectively. Parameter values *P*_*v*_ = 0.1, VE = 0.4, NI = 0.2, *m*_*v*_ = 0, γ = 73.

**Fig. A2.16:**
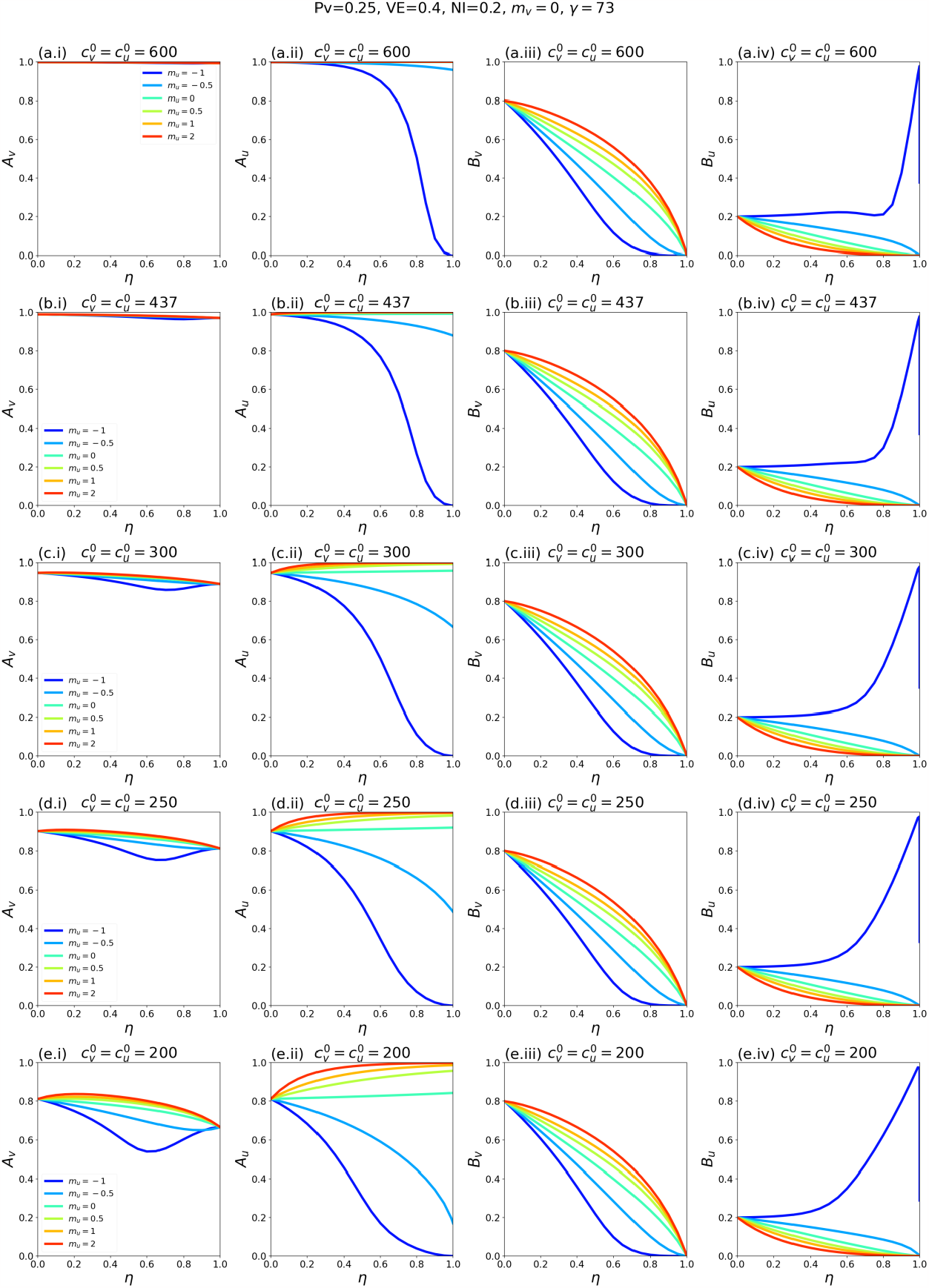
Same as Fig. A2.15, except that *P*_*v*_ = 0.25.

**Fig. A2.17:**
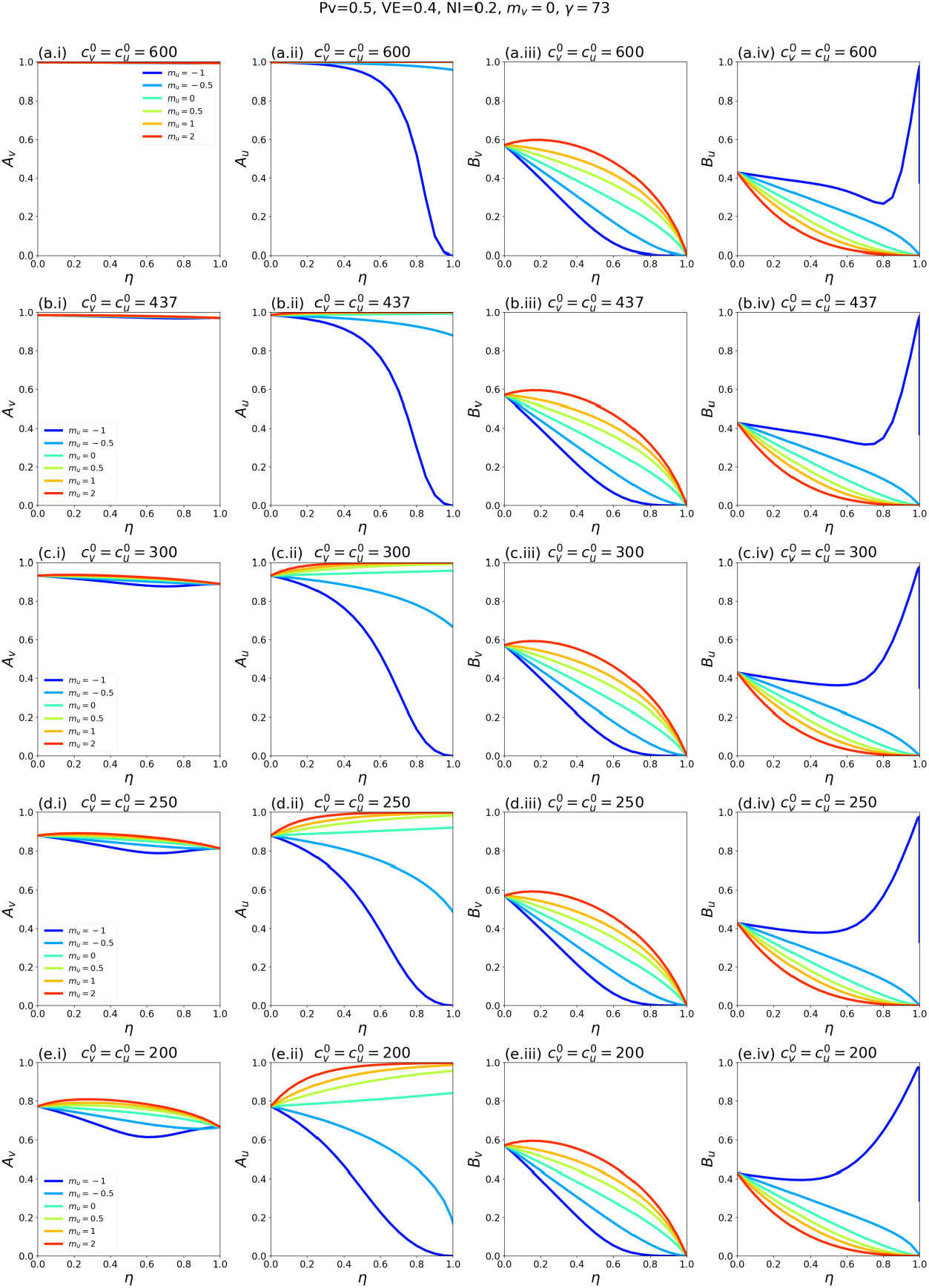
Same as Fig. A2.15, except that *P*_*v*_ = 0.5.

**Fig. A2.18:**
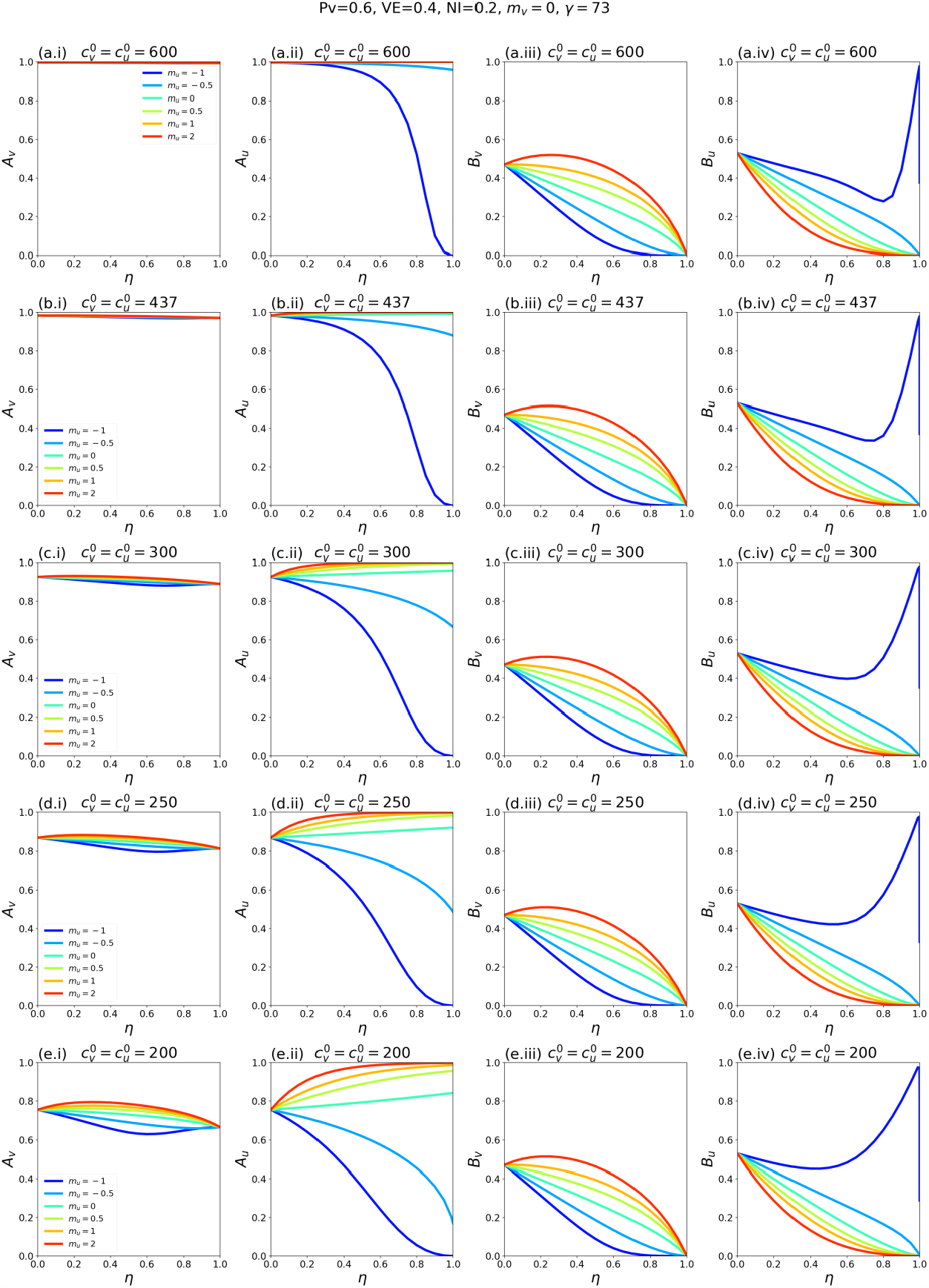
Same as Fig. A2.15, except that *P*_*v*_ = 0.6.

**Fig. A2.19:**
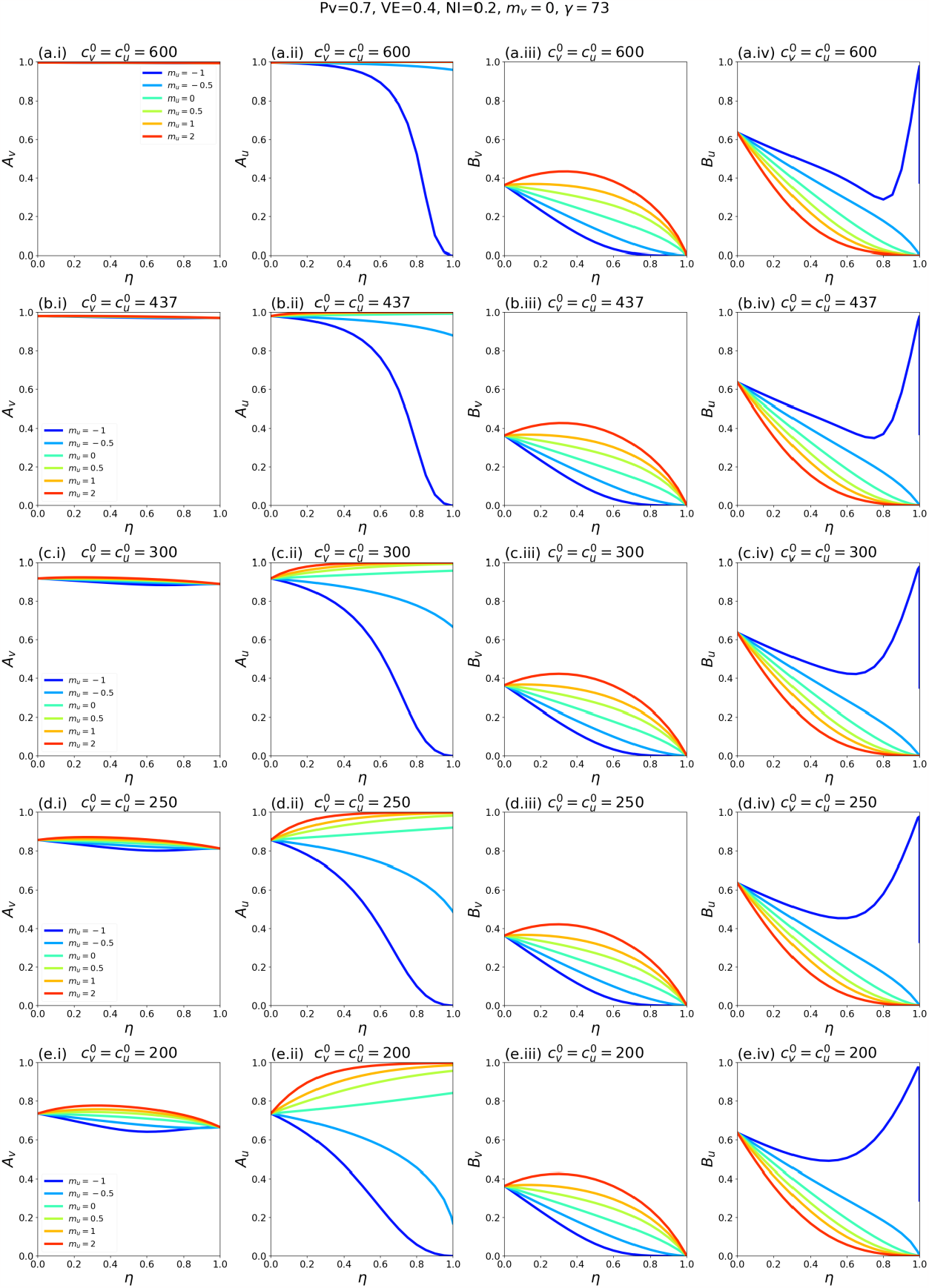
Same as Fig. A2.15, except that *P*_*v*_ = 0.7.

**Fig. A2.20:**
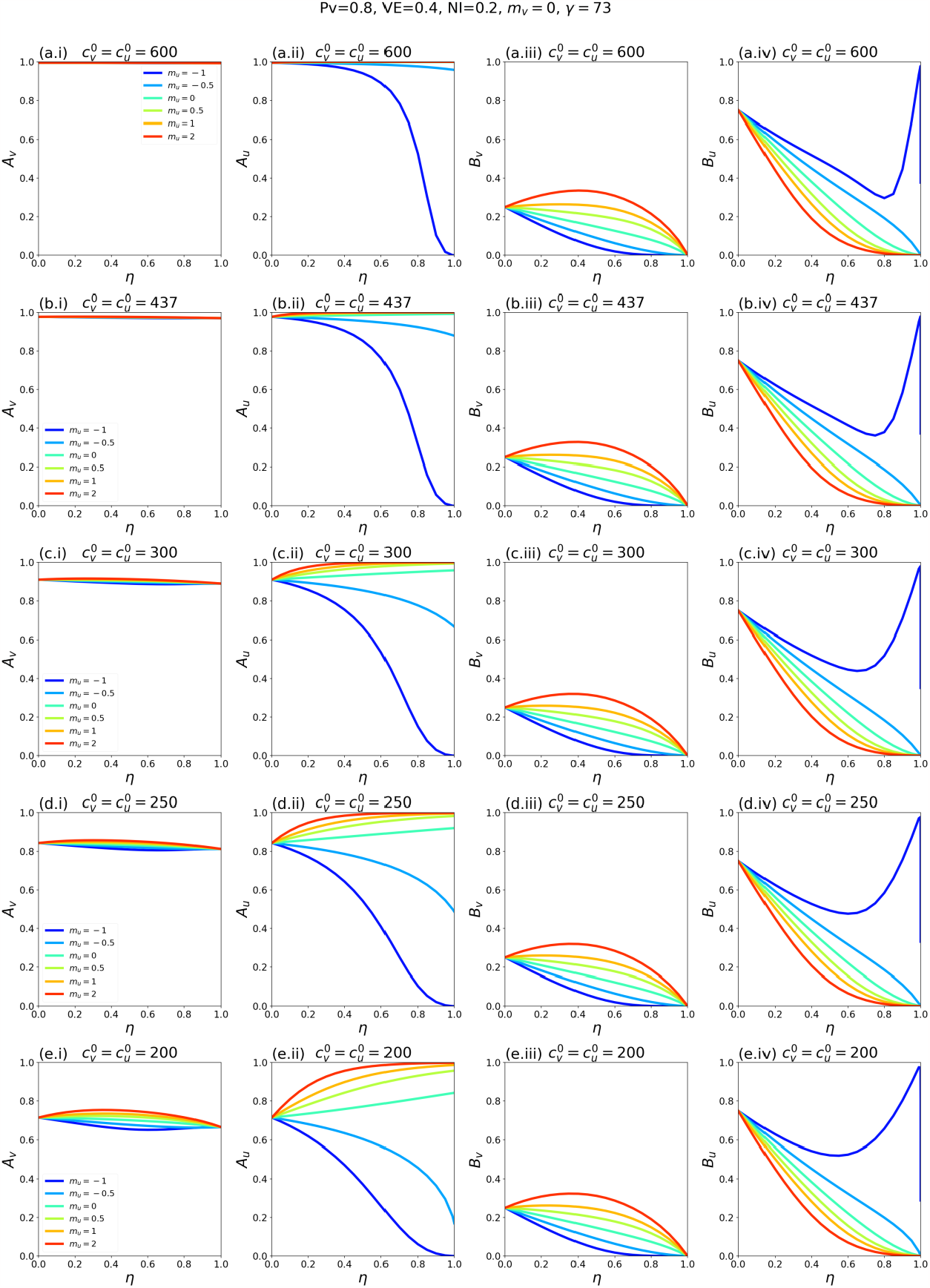
Same as Fig. A2.15, except that *P*_*v*_ = 0.8.

**Fig. A2.21:**
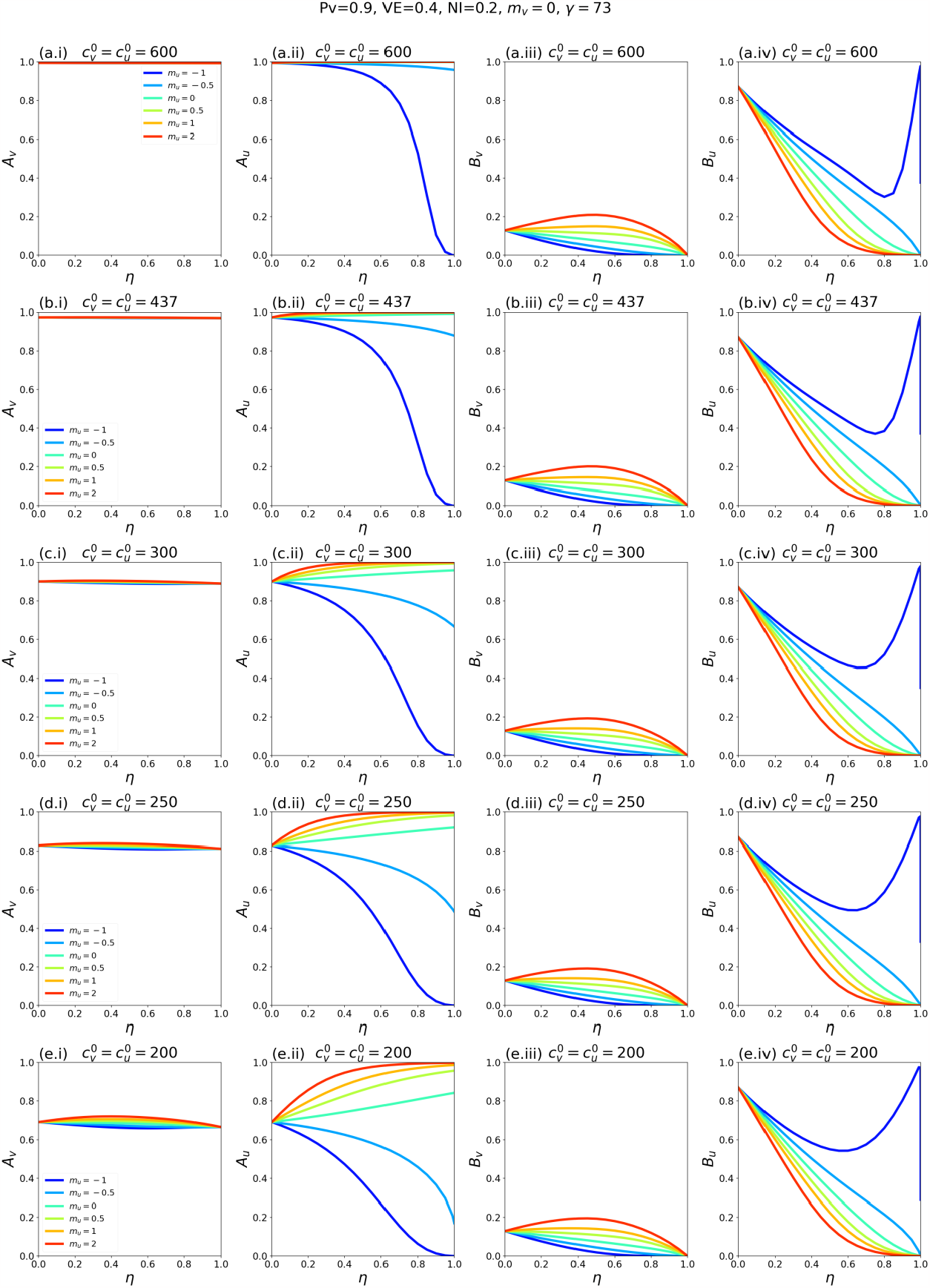
Same as Fig. A2.15, except that *P*_*v*_ = 0.9.

**Fig. A2.22:**
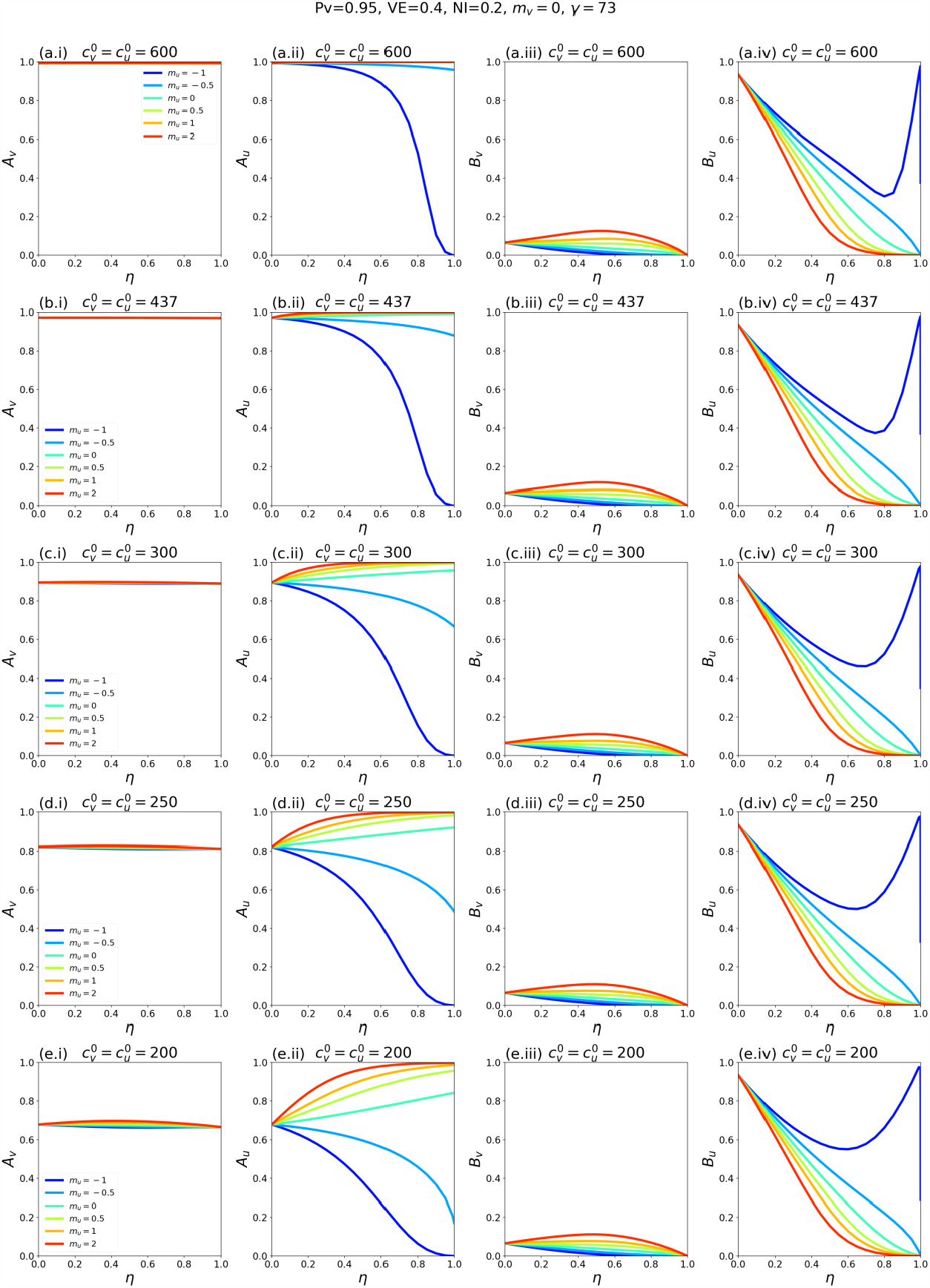
Same as Fig. A2.15, except that *P*_*v*_ = 0.95.

**Fig. A2.23:**
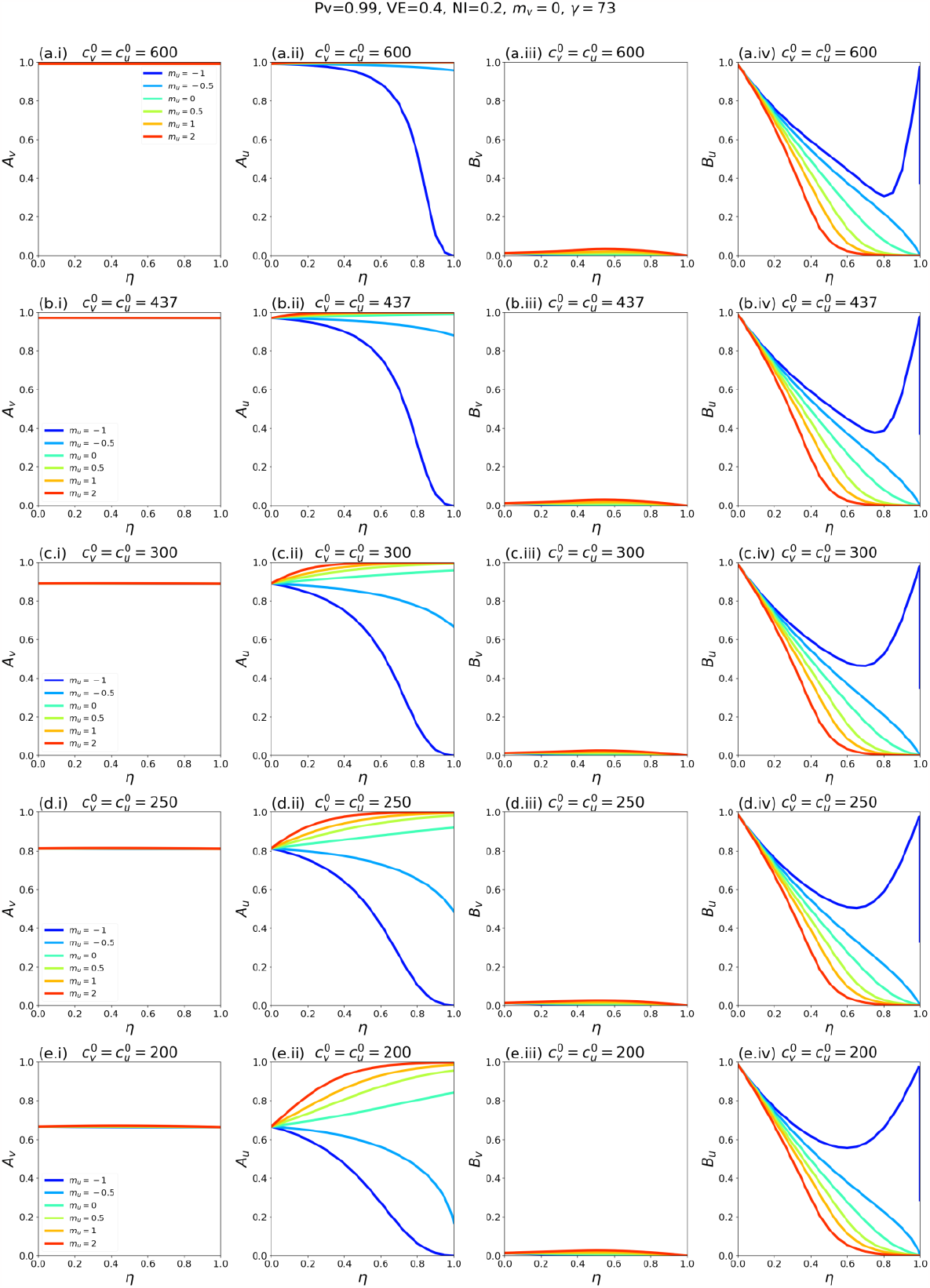
Same as Fig. A2.15, except that *P*_*v*_ = 0.99.

#### A2.3: Epidemic outcomes for different values of *m*_*v*_

**Fig. A2.24:**
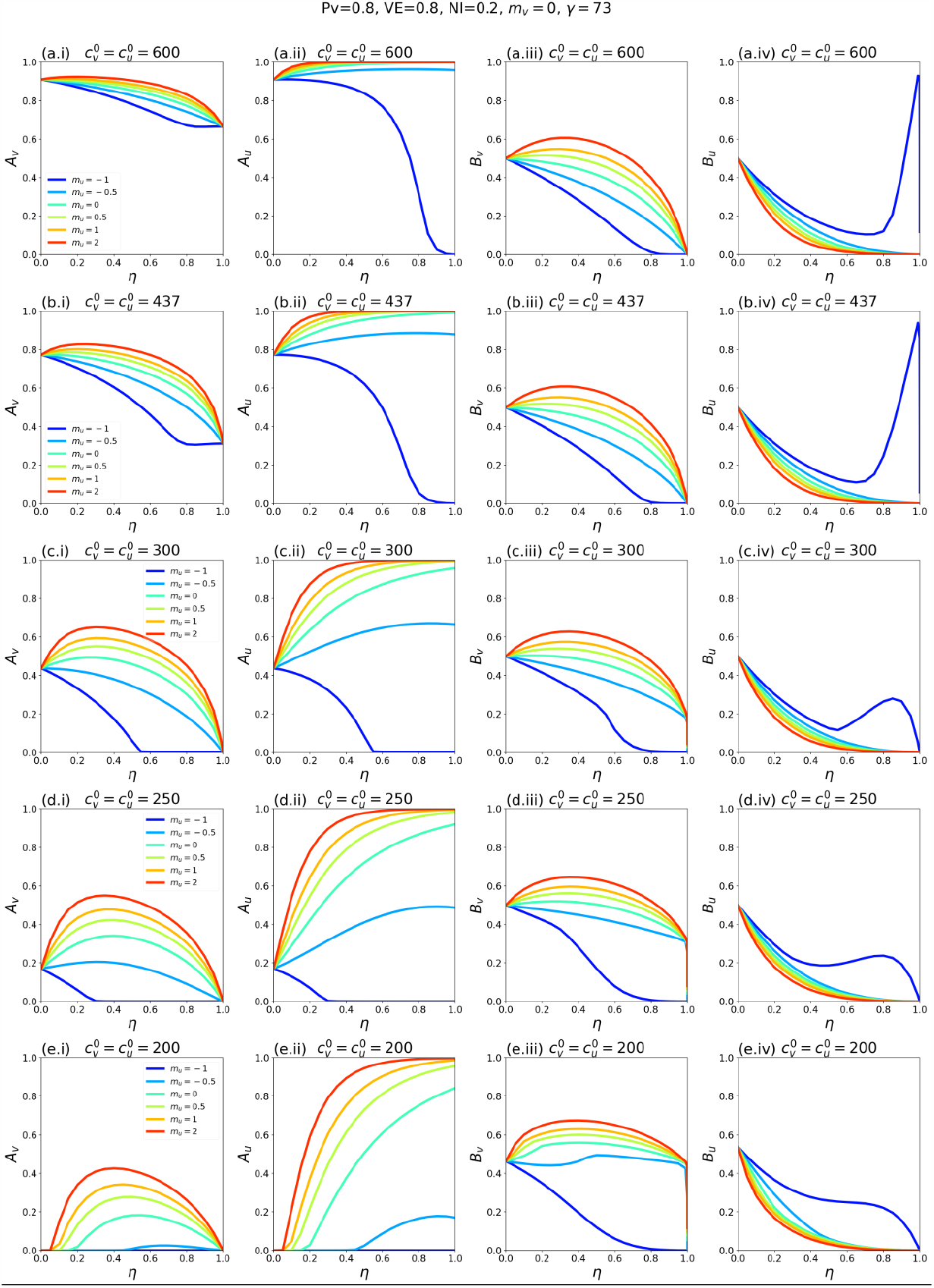
*A*_*v*_, *A*_*u*_, *B*_*v*_, and *B*_*u*_ as functions of η. Each row of panels corresponds to a choice of 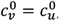, and each coloured line to a choice of *m*_*u*_ as indicated in the legends. For reference, in a single-population (no vaccination) model, the corresponding *R*_*0*_ values for rows a-e of the figure are 8.2, 6.0, 4.1, 3.4 and 2.7, respectively. Parameter values *P*_*v*_ = 0.8, VE = 0.8, NI = 0.2, *m*_*v*_ = 0, γ = 73. This figure is for the same parameters as Fig. 1 of the main text, such that the *A*_*v*_, *A*_*u*_, and *B*_*v*_ columns in this figure are reproductions of the same columns in Fig. 1 of the main text.

**Fig. A2.25:**
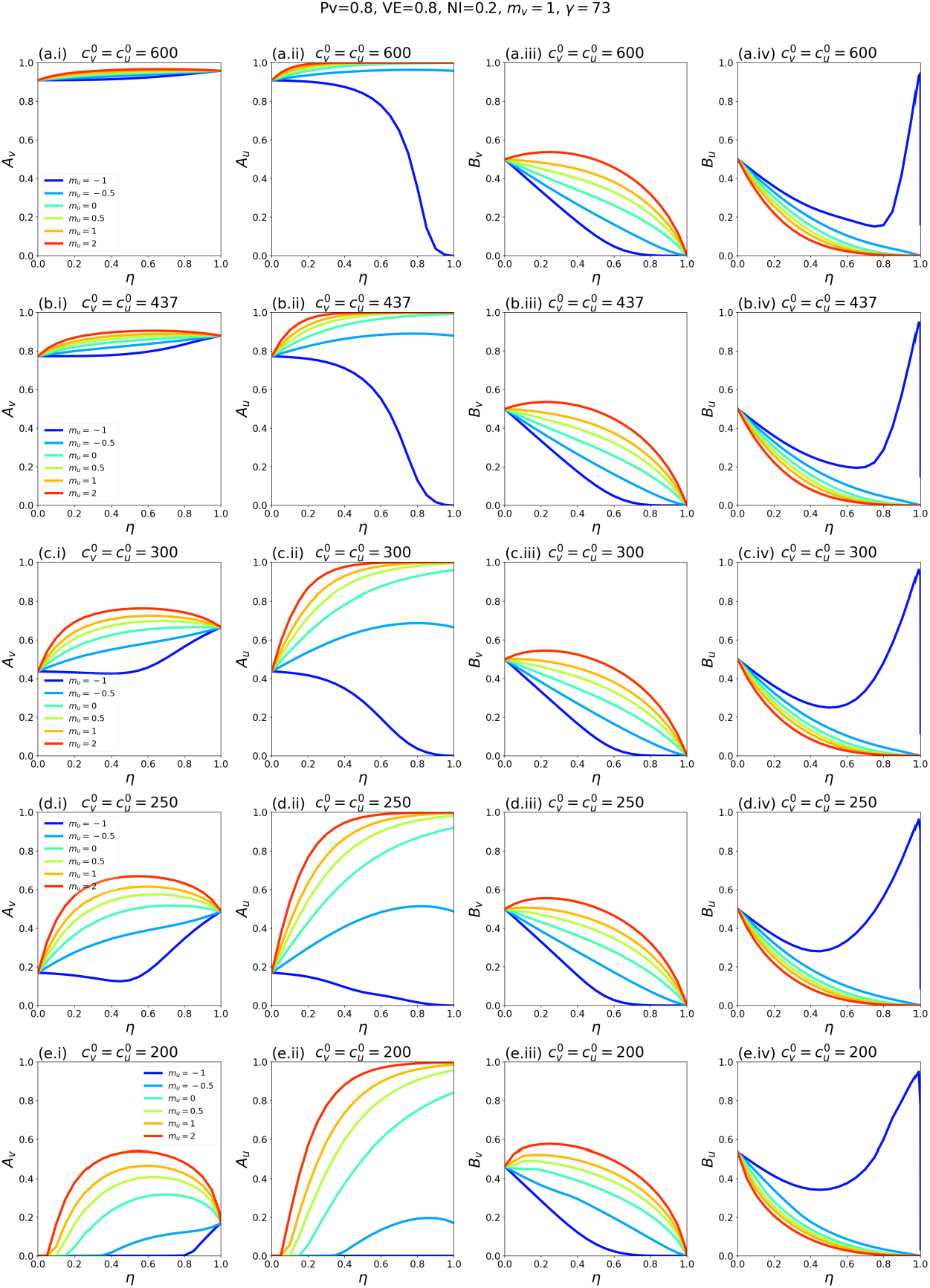
Same as Fig. A2.24, except that *m*_*v*_ = 1.

**Fig. A2.26:**
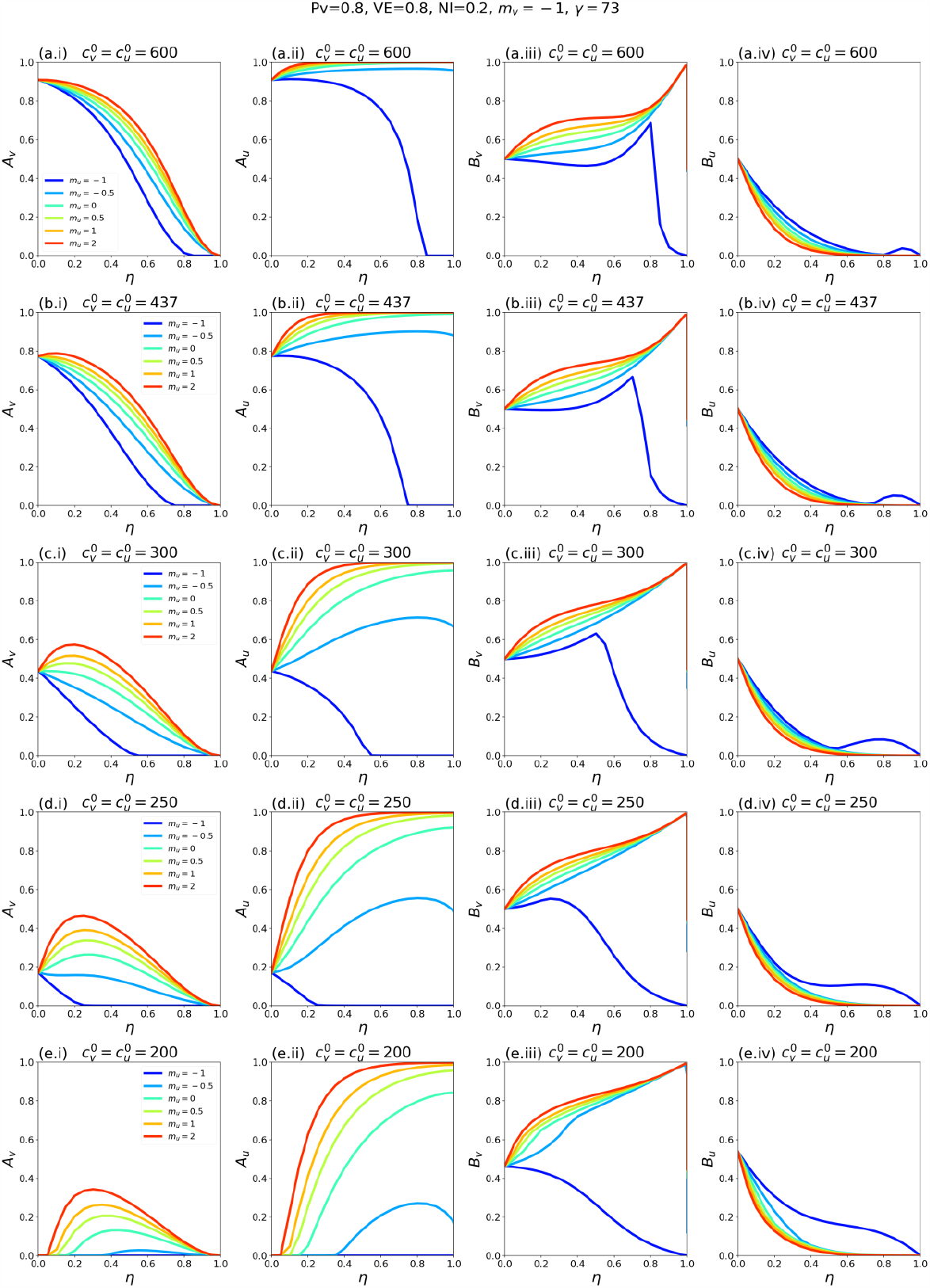
Same as Fig. A2.24, except that *m*_*v*_ = -1.

#### A2.4: Epidemic outcomes for different values of 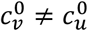

##### A2.4.1: Fixed 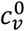, vary 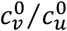

**Fig. A2.27:**
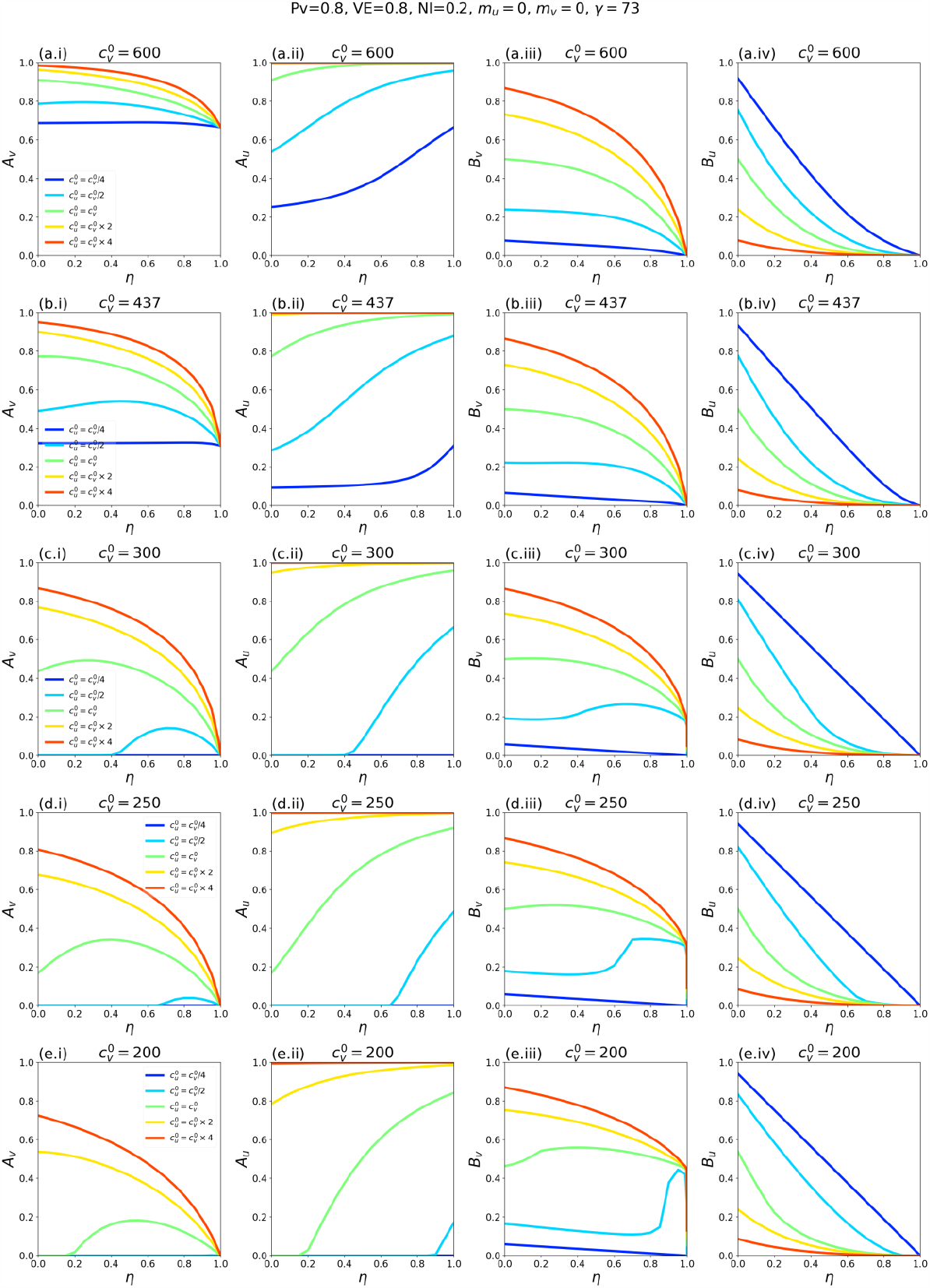
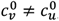, *m*_*u*_ = *m*_*v*_ = 0, γ_*u*_ = γ_*v*_ = 73, VE = 0.8, NI = 0.2, various choices of 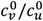 (see legend within left-column panels), showing *A*_*v*_, *A*_*u*_, *B*_*v*_,, and *B*_*u*_ as functions of η. 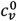 is fixed for each row in the figure and decreases moving down the rows.

**Fig. A2.28:**
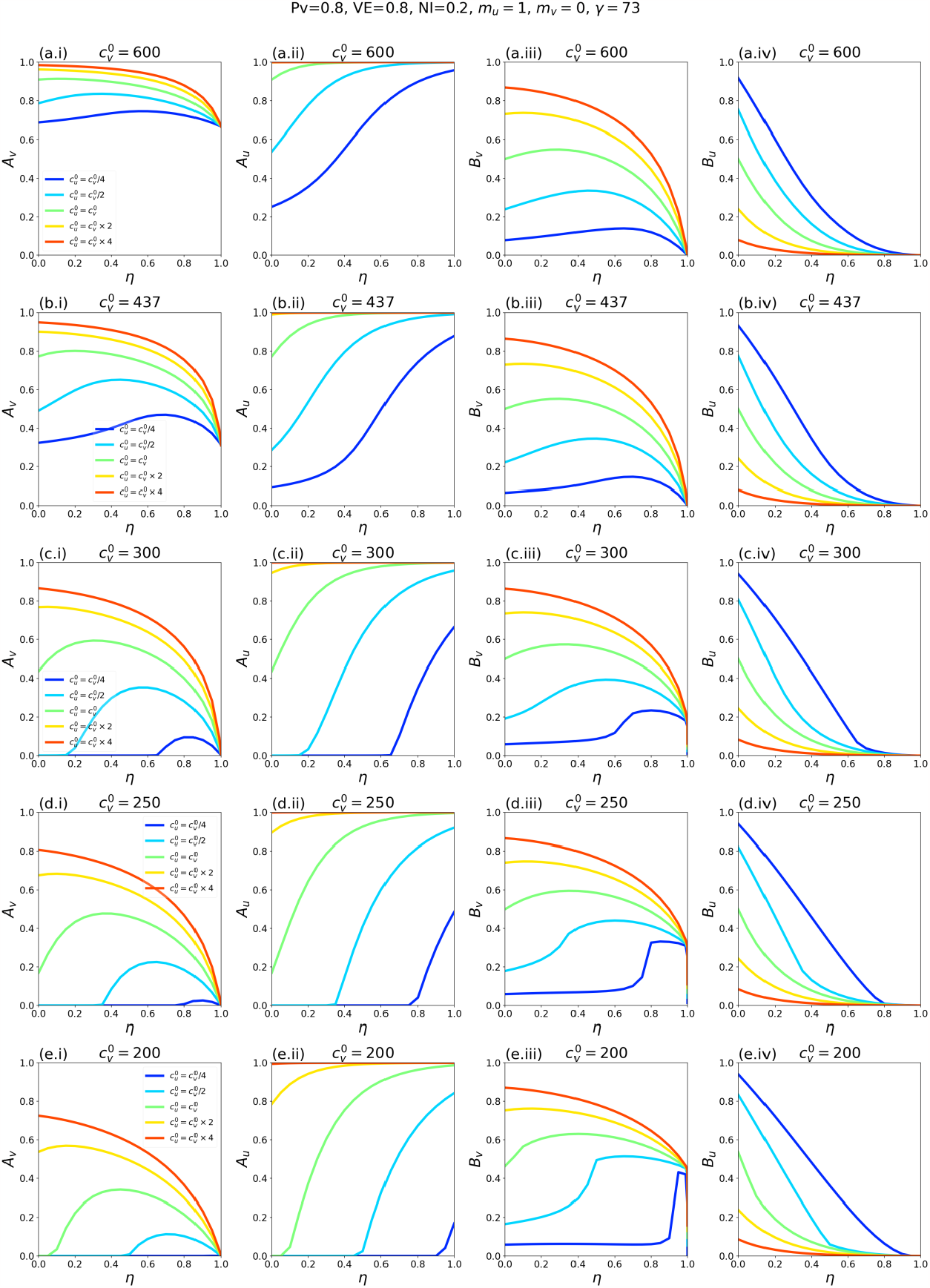
Same as Fig. A2.27, except that *m*_*u*_ = 1.

**Fig. A2.29:**
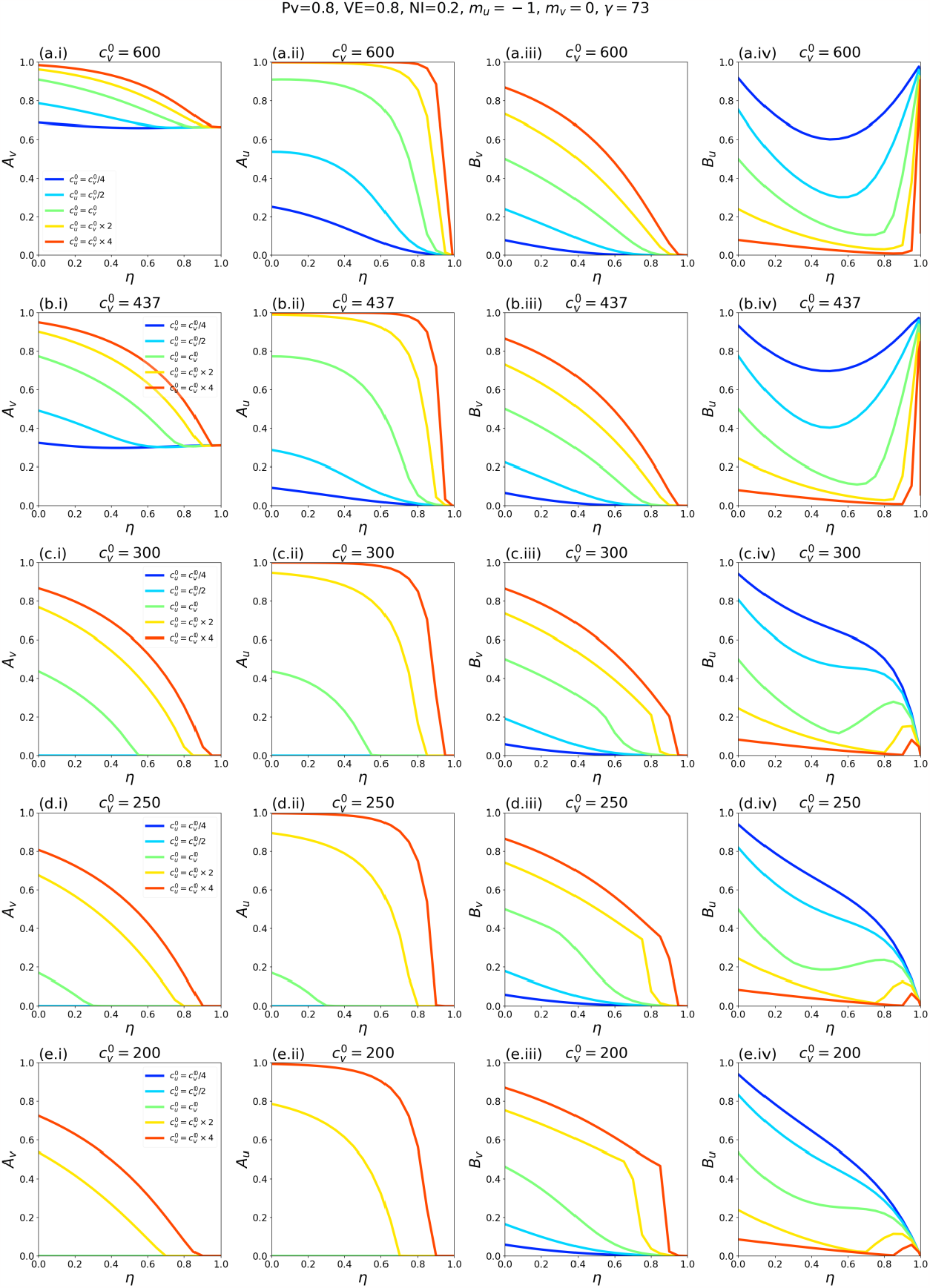
Same as Fig. A2.27, except that *m*_*u*_ = -1.

##### A2.4.2: Fixed weighted sum 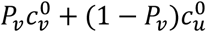

**Fig. A.30:**
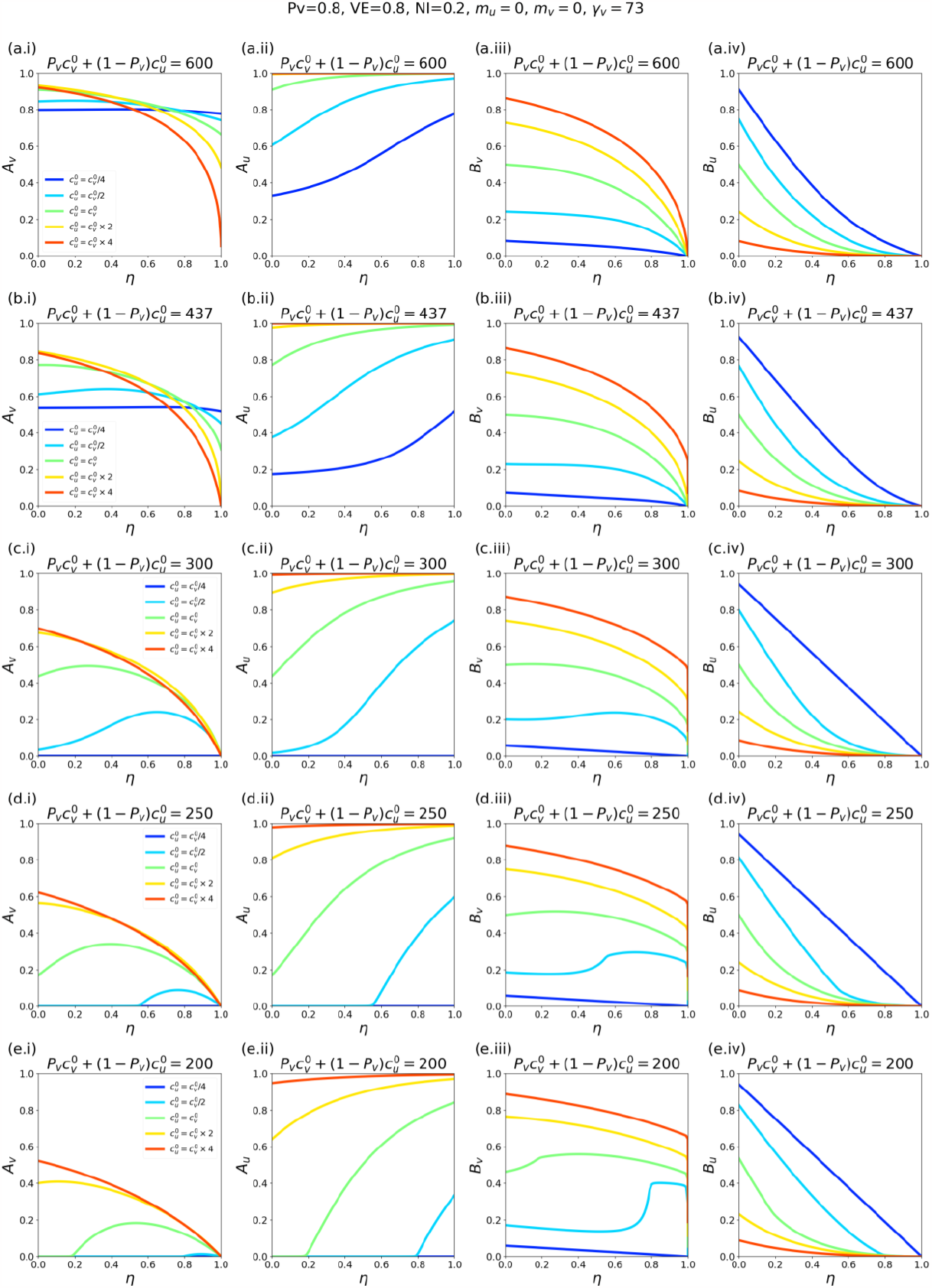
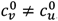, *m*_*u*_ = *m*_*v*_ = 0, γ_*u*_ = γ_*v*_ = 73, VE = 0.8, NI = 0.2, various choices of 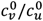 (see legend within left-column panels), showing *A*_*v*_, *A*_*u*_, *B*_*v*_,, and *B*_*u*_ as functions of η. The weighted sum 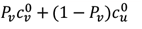 is fixed for each row in the figure and decreases moving down the rows.

**Fig. A.31:**
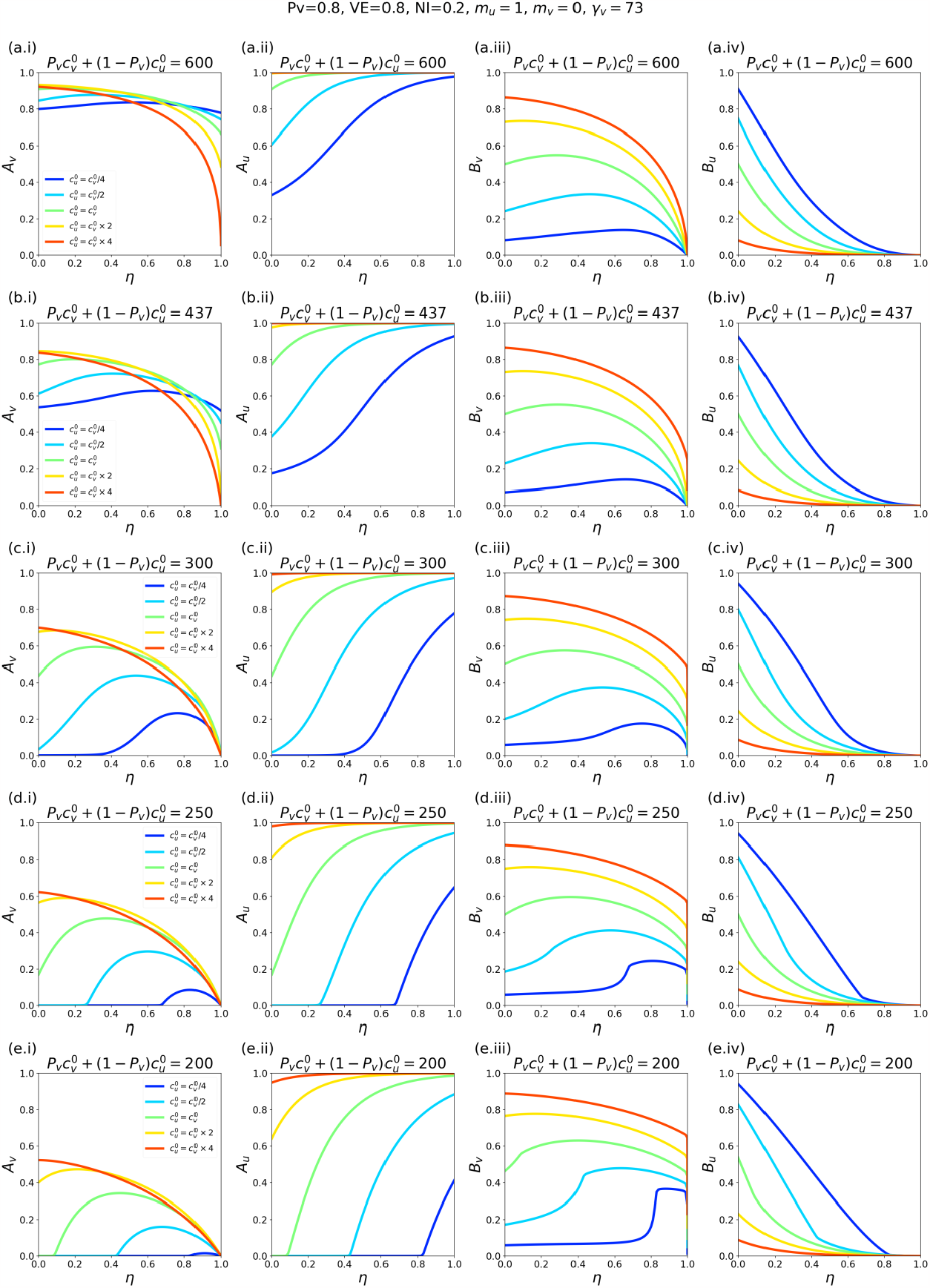
Same as Fig. A.30, except that *m*_*u*_ = 1.

**Fig. A.32:**
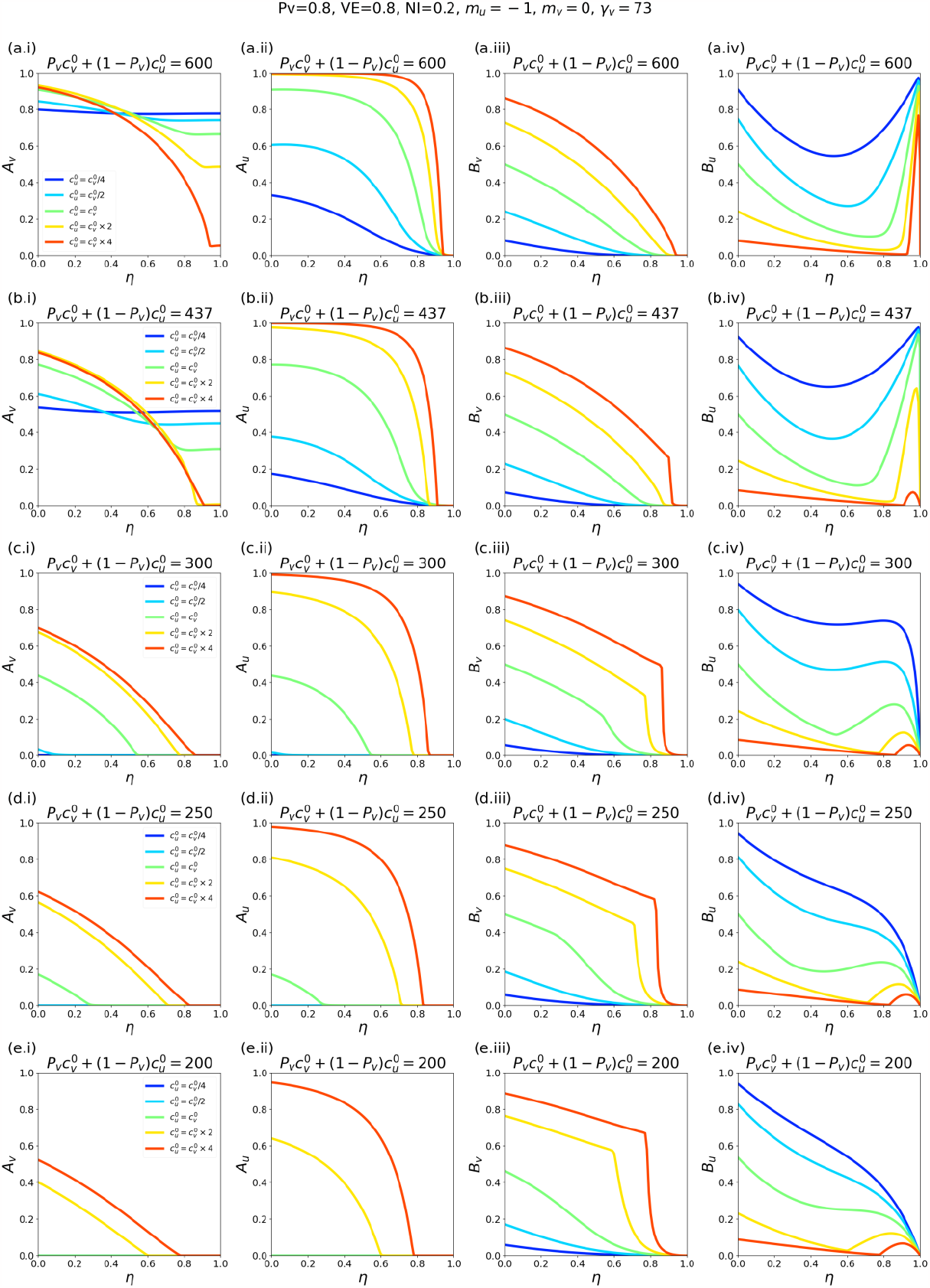
Same as Fig. A.30, except that *m*_*u*_ = -1.

## Notes

### Competing Interest Statement

The authors have declared no competing interest.

### Funding Statement

This study did not receive any funding.

### Summary of Updates

Formatting, new references, adjustments to the text, new title.

